# Assessing the Impact of Olfactory Dysfunction on Eating Behavior: A Systematic Scoping Review and Call for Standardized Assessments

**DOI:** 10.1101/2024.04.15.24305547

**Authors:** Parvaneh Parvin, Sanne Boesveldt, Elbrich M. Postma

## Abstract

Olfaction plays a priming role in both the anticipation and consumption phases of eating behavior. Olfactory dysfunction can therefore lead to changes in various aspects of eating behavior, such as food choice, appetite, and food intake. In light of the increasing prevalence of persistent olfactory dysfunction among patients affected by Covid-19, providing proper care and dietary advice to individuals with olfactory dysfunction is imperative. Therefore, this scoping review seeks to gain a better understanding of the impact of olfactory dysfunction on eating behavior. Following the PRISMA guidelines, 49 papers were included, the outcomes were presented by dividing them into two categories: 1) anticipatory eating behavior, including (anticipatory) food liking, appetite and craving, food preferences, food neophobia, and cooking habits; and 2) consummatory behavior, including, food intake, consumption frequency, adherence to dietary guidelines, (experienced) food liking, food enjoyment, and eating habits. Our results show that in the anticipatory phase of eating behavior, food liking and, food preferences and in the consummatory phase, food enjoyment is most affected in people who experienced a sudden change in olfactory function rather than a gradual decline. Moreover, changes in food flavor perception due to olfactory dysfunction, result in a shift of food preferences towards more taste-based preferences, such as salty or savory (i.e., umami) foods. Subsequently, changes in preferences can affect food intake and adherence to dietary guidelines, but only to a limited extent. Appetite is more likely to be low in individuals with short-term olfactory dysfunction compared to those with long-term changes. Generally, eating behavior is more impacted in individuals with a distorted sense of smell than in those with smell loss, and the effect becomes more pronounced over time. Due to the heterogeneity of methods used to measure different aspects of eating behavior, this review stresses the importance of more research on olfaction and eating behavior using standardized and validated assessments. Such research is essential to better understand the effects of olfactory dysfunction on each aspect of eating behavior and provide effective interventions.

**Highlights:**

- Food liking, preferences, and enjoyment are the most affected by olfactory dysfunction
- Impact on eating behavior is more pronounced in qualitative vs. quantitative smell loss
- Effects of olfactory dysfunction vary by duration and nature (qualitative vs. quantitative)
- Standardized, validated methods are needed to assess eating behavior in future studies
- There is a crucial need for effective interventions to enhance the eating experience

**Graphical Abstract:** 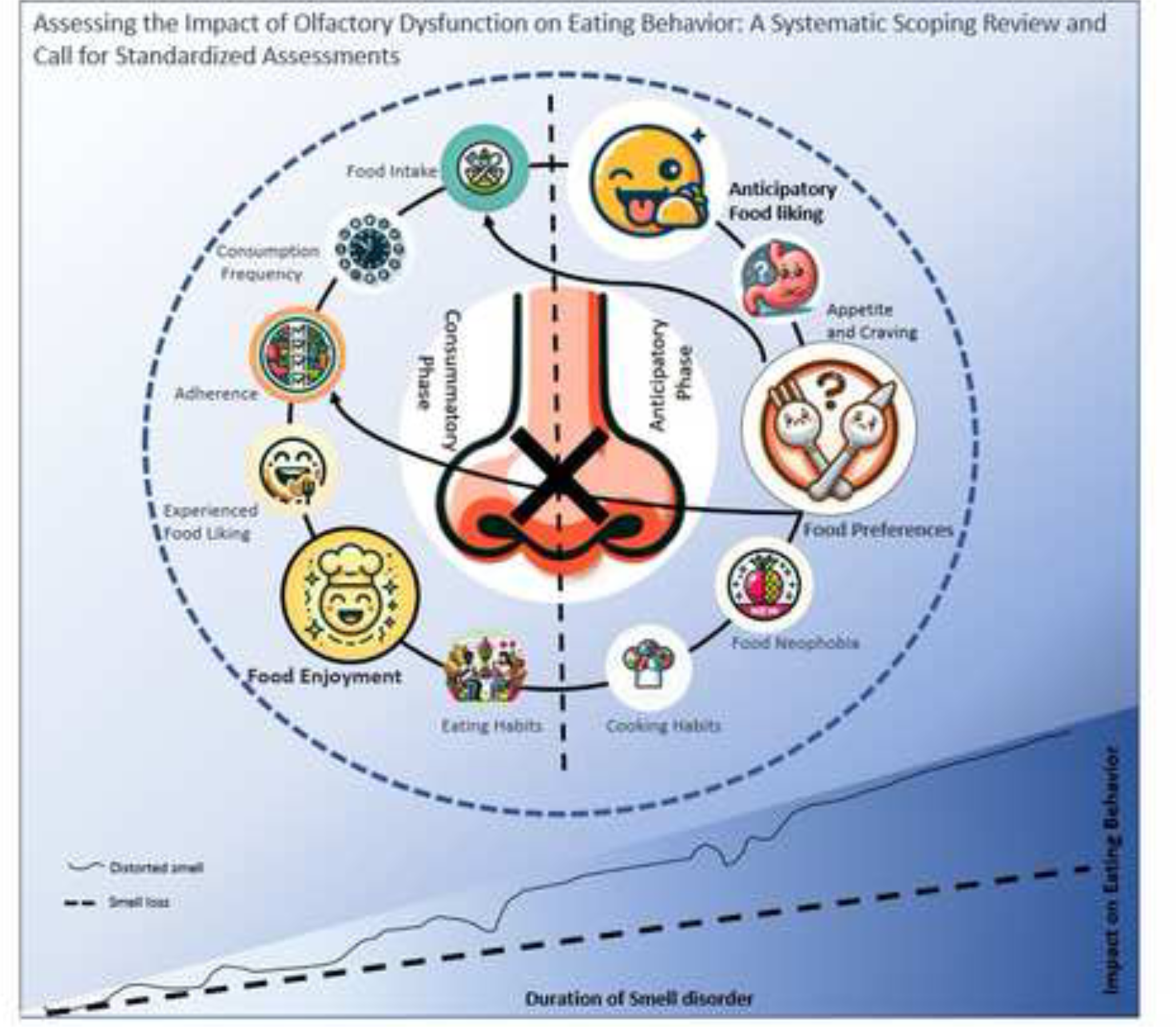

## Introduction

Olfaction is a crucial sense that plays a pivotal role in several essential functions in our daily lives. It assists in social communication, facilitates avoidance of environmental hazards, and contributes to eating behavior, such as the enjoyment of food (Aschenbrenner et al., 2008). Olfactory dysfunction has been linked to various disturbances in the three aforementioned areas, incorporating social issues related to hygiene and altered sexual behaviors (Blomkvist & Hofer, 2021), difficulty detecting hazardous smells or food, and reduced food enjoyment (Stevenson, 2009). These deficits can have a profound impact on an individual’s quality of life (Boesveldt et al., 2017), including eating behavior, as olfaction is closely related to our sense of taste and is a significant factor in our perception of flavors. Olfactory dysfunction refers to a decreased ability or distorted ability to smell during sniffing (orthonasal olfaction) or eating and drinking (retronasal olfaction). It is typically divided into two categories: quantitative and qualitative dysfunction. Quantitative dysfunction can be divided into anosmia, a complete loss of sense of smell, and hyposmia, a reduced sense of smell. On the other hand, qualitative dysfunction is defined as a change in the quality of perceived odors and comprises parosmia, a distorted sense of smell, and phantosmia, odor hallucinations. The major difference between these two qualitative disorders is that these distorted olfactory sensations are experienced in the presence or absence of an odor, respectively (Desai & Oppenheimer, 2021), (Leopold, 2002).

It has been estimated that between 3-20% of the general population experiences either qualitative or quantitative olfactory dysfunctions (Boesveldt et al., 2017), (B. N. Landis & Hummel, 2006). Up to 1% of those individuals may have a congenital form, such as Kallmann’s syndrome or isolated congenital anosmia (Temmel et al., 2002). Other causes for olfactory dysfunction include (1) head trauma, (2) other viral infections such as influenza (Soler et al., 2020), (3) nasal causes such as sinusitis or polyposis nasi, (4) aging and (5) age-related neurological illnesses such as Parkinson’s and Alzheimer’s disease (Hummel et al., n.d.). Recently, COVID-19 has been linked to persistent loss and alteration of smell, leading to an increase in the number of individuals experiencing olfactory dysfunction (Soler et al., 2020), (Boscolo-Rizzo et al., 2020), (Hopkins & Kelly, 2021).

To measure olfactory dysfunction, both subjective and objective measures can be used. Subjective measures of smell are typically questionnaires that ask individuals to rate their level of olfactory ability and can be completed in a person’s own home or online. Objective measures, on the other hand, are mostly carried out under the supervision of a researcher or healthcare provider and typically consist of psychophysical tests developed to measure and quantify human responses to physical stimuli (Hannum et al., 2020), like the Sniffin’ Sticks test (Oleszkiewicz et al., 2019) or the UPSIT (Doty et al., 1984). These tests can assess distinct aspects of olfactory function, such as the ability to detect, identify or discriminate odors.

Eating behavior is a multifaceted process that is influenced by physiological, psychological, and behavioral factors elicited by the sensory and nutritional properties of foods (Blundell et al., 2010). This multifaceted process can be divided broadly into two distinct phases: *anticipatory* and *consummatory phases of eating behavior*. In the anticipatory phase, which encompasses the pre-ingestive aspect of eating, the body prepares for eating by secreting hormones that increase hunger and stimulate the digestive system. In this phase, sensory cues, such as (orthonasal) ambient food odors, can have a substantial influence on appetite, cravings, food preferences, and influence decisions regarding food choice and consequently the food consumption (Ramaekers et al., 2016). Additionally, food liking, but also food neophobia, the reluctance or fear of trying new or unfamiliar foods, shapes individuals’ food choices and preferences. The anticipatory phase also comprises cooking habits, including food preparation. Upon transitioning to the consummatory phase of eating behavior, the focus shifts to the actual consumption (ingestive) process. In this phase, retronasal odor, but also taste, and texture play a crucial role in overall flavor perception and determining the amount and type of food that is consumed. These factors contribute to food intake behavior including nutritional intake, consumption frequency, and adherence to dietary guidelines. Moreover, the pleasure derived from consumed food during the consummatory phase is paramount, as it influences individuals’ perception of the meal and subsequent dietary choices. Long-term eating habits, such as behaviors and routines individuals have when it comes to consuming food and the social aspects of eating, also play a crucial role in shaping individuals’ overall consumption patterns, and meal planning (Aschenbrenner et al., 2008).

When measuring eating behavior, the choice of techniques employed depends on the research purpose, study design, and the specific aspect of eating behavior being evaluated, such as food preference or food intake. These methods range from simple visual analog ratings of liking or appetite to more elaborate questionnaires or (behavioral) tasks to assess preferences, to food frequency questionnaires and food intake diaries (Dovey, 2011). Additionally, researchers must choose between laboratory-based studies and free-living studies to measure eating behavior, which often leads to a focus on specific subsets of eating behavior aspects.

While the importance of olfaction in eating behavior is well-established, the impact of olfactory dysfunction on distinct aspects of eating behavior is not yet fully understood. Thus, this scoping review sought to identify and summarize evidence on the impact of olfactory dysfunction on distinct aspects of eating behavior as alluded to above. By examining the effects of olfactory dysfunction on these various facets of eating behavior, this review aims to provide a comprehensive understanding of how olfactory impairment impacts individuals’ overall eating behavior.

## 2. Materials and Methods

### 2.1 Literature search strategy

The literature search and screening were performed following the Preferred Reporting Items for Systematic Reviews and Meta-Analyses guidelines (PRISMA) (Moher et al., n.d.), as shown in Figure 1. The search covered papers on human studies published between the earliest record and July 2022. This search was executed on the Scopus, Ovid MEDLINE, PubMed, and PsycINFO databases, chosen for their relevance and comprehensive coverage in the field. The general search strategy involved searching for title, abstract, and keywords (or headwords on the PsycINFO database). The specific combinations of keywords used are detailed in Table 1, while database-specific search strategies can be found in Appendix 1. Results from each database are tabulated in Table 2.a. Additionally, a manual search in the reference list of the articles included was performed to identify further eligible studies. It’s worth noting that we expanded our search criteria later in the process, as detailed in sections 2.1.1 and 2.2.1. The result of the additional search is presented in Table 2.b.

**Figure 1.**
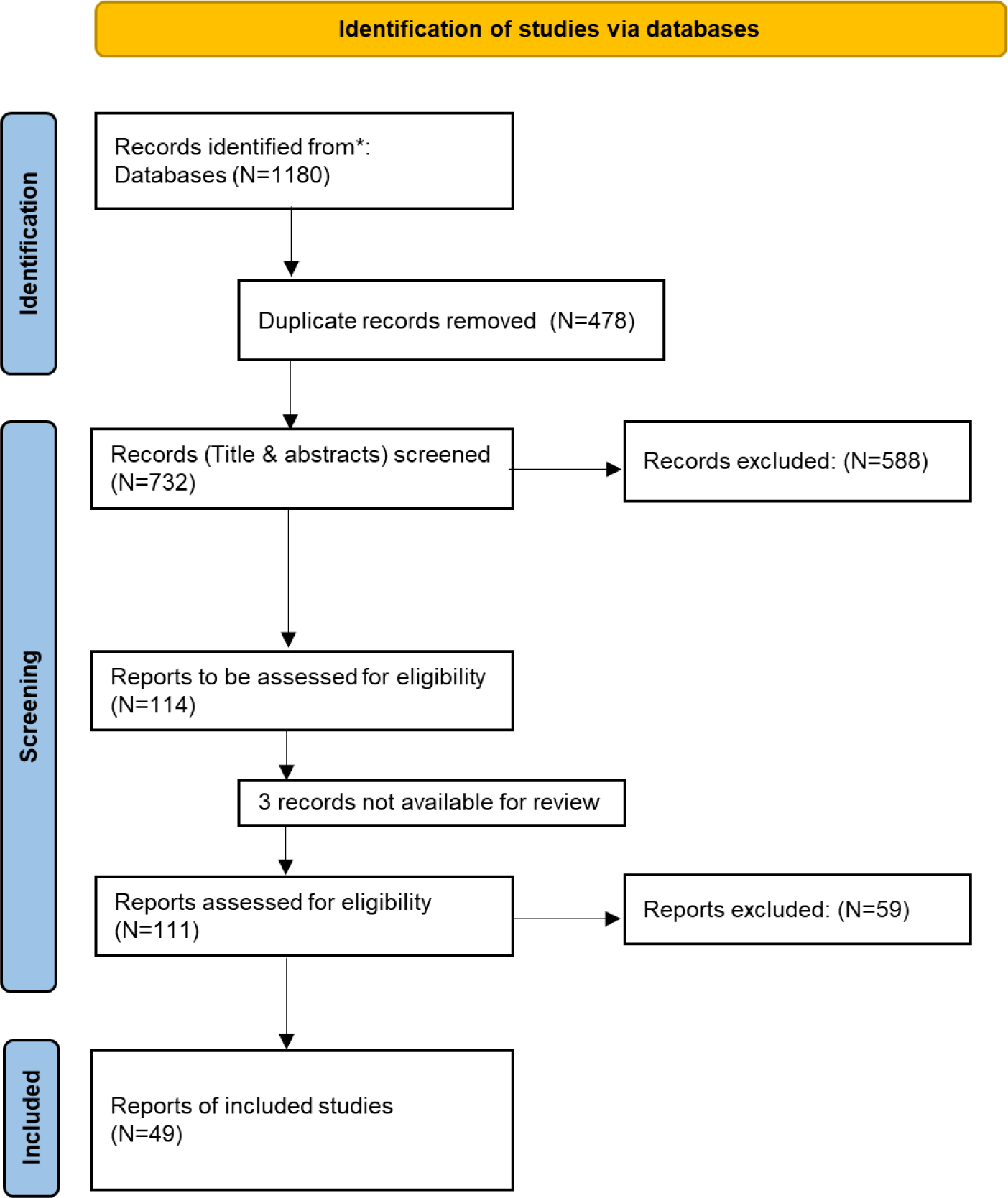
Flow chart showing the results of the literature search on studies investigating the effect of olfactory dysfunction on eating behavior.

**Figure 2:**
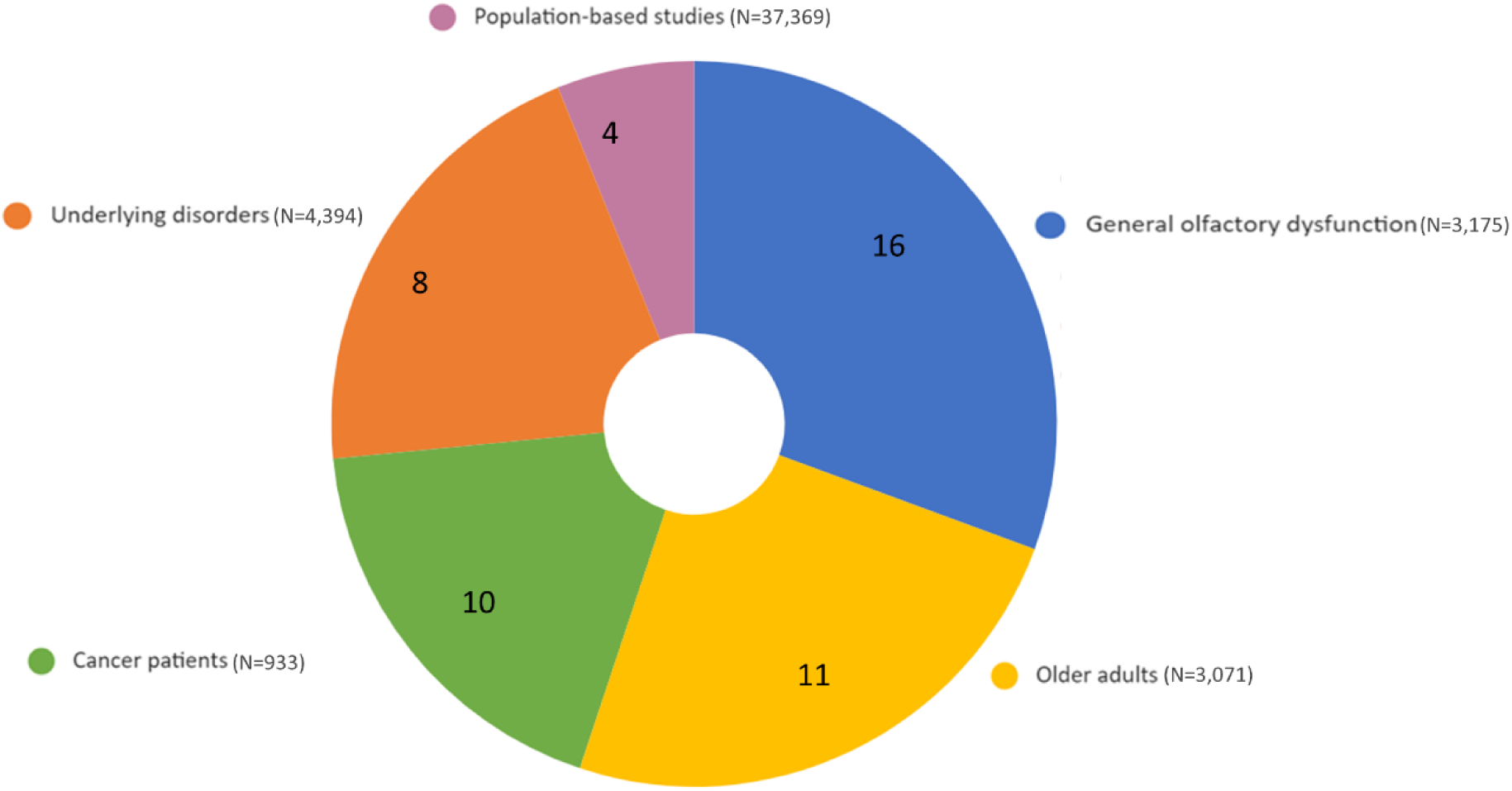
Distribution of populations across studies (Number of Studies by Category with Participant Counts)

**Table 1.** Main keywords for olfactory dysfunction and eating behavior used for the literature search.

| Keywords | Strings and combinations of keywords |
| --- | --- |
| Olfactory Dysfunction | (( olfact* OR smell OR chemosensory OR odor ) W/5 ( disorder OR loss OR dysfunction OR changes OR deficit OR impairment OR decrease OR alter* )) OR TITLE-ABS-KEY ( ( anosmia OR hyposmia OR parosmia ) ) AND |
| Eating behavior | appetite OR diet* OR food OR eating OR feed* ) W/5 ( habit OR intake OR pattern OR preference OR pleasantness OR liking OR wanting OR neophobia OR choice OR enjoyment OR perception OR behavi* OR pleasure OR quality OR consumption) |

**Table 2.a.** Search results per database for the initial literature search.

| Database | Original # of Items found | # of deleted duplicates | # of unique items found |
| --- | --- | --- | --- |
| Medline® | 237 | 22 | 215 |
| PubMed | 265 | 98 | 167 |
| PsycINFO® | 95 | 5 | 90 |
| Scopus | 463 | 147 | 316 |
| Manual search | 2 | 0 | 2 |
| Check for external duplicates (all databases in one) | 0 | 132 | -132 |
| Total | 1062 | 404 | 658 |

**Table 2.b.**
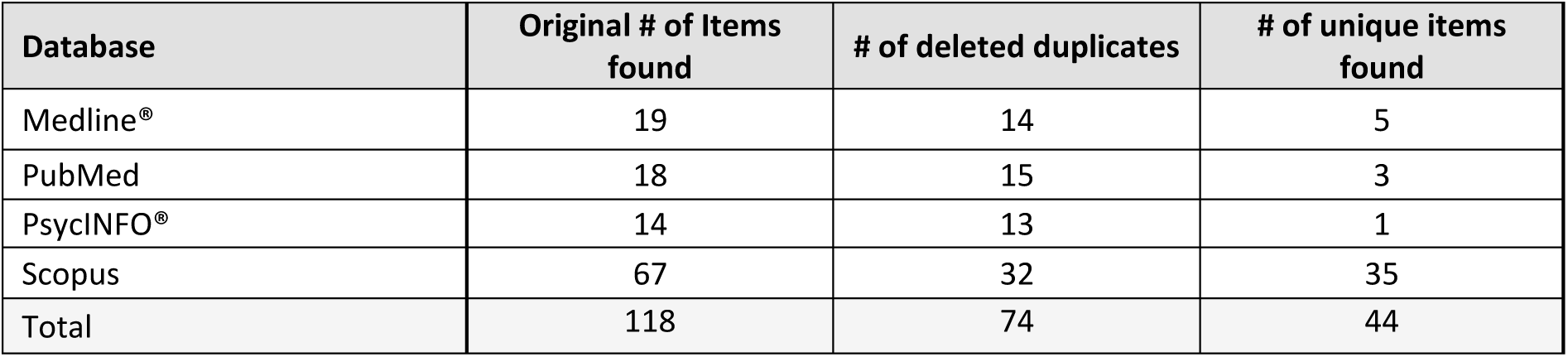
Search results per database for the additional literature search.

| Database | Original # of Items found | # of deleted duplicates | # of unique items found |
| --- | --- | --- | --- |
| Medline® | 19 | 14 | 5 |
| PubMed | 18 | 15 | 3 |
| PsycINFO® | 14 | 13 | 1 |
| Scopus | 67 | 32 | 35 |
| Total | 118 | 74 | 44 |

#### 2.1.1 Extended Literature Search

At the outset of our scoping review, we conducted an initial search focusing on particular aspects of eating behavior. These aspects were chosen due to their well-established status as domains of eating behavior extensively explored in the literature. However, as we reviewed the final selection of papers, it became evident that additional terms, such as food liking, (food wanting), and food neophobia, were identified as significant factors that had not been initially included in our categorization. We acknowledged the significance of these factors, particularly concerning olfactory dysfunction and its impact on eating behavior. As a result, we broadened our search strategy to include these new terms in our scoping review, using the same conditions as mentioned in Section 2.1.

### 2.2 Study selection process

In the initial search, in total, 1062 results (see Table 2.a) were identified both using search strategies (N=1060) and through reference lists (N=2). After removing 404 duplicate records, the remaining 658 articles were assessed for eligibility by two independent reviewers (PP and EP) based on title and abstract. First, a selection of 40 articles was screened by both reviewers, and Cohen’s kappa was calculated. The inter-rater agreement for the exclusion of articles was high (K = 0.93; almost perfect agreement above 0.80 (J. R. Landis & Koch, 1977)). Next, both reviewers screened titles and abstracts of all 658 articles, using the following exclusion criteria: animal studies; no eating behavior; no olfactory dysfunction; case study; no original research article; not related to the research question (i.e., the effect of olfactory dysfunction on eating behavior). We excluded studies on eating disorders as ‘not related to the research question’, as our primary focus was to investigate healthy eating behavior. Therefore, studies that specifically examined disordered eating patterns or individuals with diagnosed eating disorders were not included. See Table 3.a, for a list of excluded articles.

**Table 3.a.** Results from the title and abstract screening for the initial search.

| Exclusion criteria | Number of papers excluded |
| --- | --- |
| Animal study | 17 |
| No eating behavior | 97 |
| No olfactory dysfunction | 193 |
| No original research article | 176 |
| Not related to the research question | 35 |
| case study | 22 |
| duplicated | 7 |
| Total number of papers excluded | 547 |

As the last step, both reviewers performed full-text screening for all articles that passed the abstract screening. In total, 108 full texts were reviewed, using the same exclusion criteria as used during the title and abstract screening, with ‘no English full text available’ added as an additional criterion. After review, 61 articles were excluded, resulting in 47 full-text articles for quality assessment and data extraction.

### 2.3 Additional literature search

In total, 118 articles were found, of which 44 new articles were identified after the removal of the duplicate articles from our previous search. The articles underwent the procedures outlined in Section 2.2. Following the title and abstract screening, 3 papers advanced to full-text screening and were included in the final selection for quality assessment and data extraction. See Table 3.b for a list of excluded articles.

**Table 3.b.** Results from the title and abstract screening for additional search.

| Exclusion criteria | Number of papers excluded |
| --- | --- |
| Animal study | 6 |
| No eating behavior | 2 |
| No olfactory dysfunction | 3 |
| No original research article | 5 |
| Not related to the research question | 24 |
| Not available in English | 1 |
| Total number of papers excluded | 41 |

### 2.4 Quality assessment and data extraction

Quality assessment was done using the Quality Assessment Tool for Observational Cohort and Cross-Sectional Studies from the National Heart, Lung, and Blood Institute (*Study Quality Assessment Tools | NHLBI, NIH*, n.d.). This tool contained 14 questions on the paper’s quality that were answered for all included full texts using Yes, No, Cannot Determine, Not Applicable, or Not Reported (see Table A2.1 in Appendix 2).

Based on the established criteria, all papers were categorized as either Good, Fair, or Poor. Out of 47, 30 papers received a ’Good’ rating, 17 were rated as ’Fair’, and none were deemed ’Poor’. As a result, no papers were excluded based on the Quality Assessment. Papers sourced from the additional search were also subjected to a quality assessment: one paper was rated ’Good’, another as ’Fair’, and the third as ’Poor’. Consequently, the latter was excluded from the selection. A detailed breakdown of the ratings, organized by the etiology of olfactory dysfunction, can be found in Table A2.2, Appendix 2. This process culminated in a final selection of 49 papers for the comprehensive review.

For the final selection of papers, all available data were obtained regarding the first author’s name, year of publication, study design (e.g., cross-sectional or longitudinal), demographics of the participants (i.e., sample size, mean age ± SD/SEM, age range, gender, and characteristics of the study groups), type of olfactory function measures utilized (e.g., identification, discrimination, threshold, detection, self-report), eating behavior measures employed, and primary findings. Appendix 3 includes a full overview of the methods used to measure olfactory function.

### 2.5 Presentation of results

The primary findings were organized into two categories, based on the literature search findings: the anticipatory phase of eating behavior, which includes food neophobia, (anticipatory) food liking, food preferences, appetite and craving, and cooking habits; and the consummatory phase of eating behavior, which encompasses food intake, consumption frequency, adherence to dietary guidelines, (experienced) food liking, food enjoyment, and eating habits (see Table 4 and Table 5 for details). In the manuscript’s result section, within each section, findings are presented according to the underlying cause of olfactory dysfunction in the study population (i.e., general olfactory dysfunction, older adults, cancer patients, and other underlying diseases). This categorization allowed for a comprehensive examination of olfactory dysfunction across different patient populations and conditions.

**Table 4.** Included articles for the anticipatory phase of eating behavior per category (food neophobia, food liking, food preferences, and appetite and craving)

| Anticipatory phase of eating behavior |  |  |  |  |  |
| --- | --- | --- | --- | --- | --- |
|  | Participants |  | Measurement methods |  |  |
| Authors | Sample | Age: mean ± SD (age range);<br>Gender: F /M | Olfactory<br>dysfunction | Eating behavior | Result |
| **Food neophobia** |  |  |  |  |  |
| (Manesse et al., 2021) | French individuals (N=3685)<br>Older adults (N=202) (subgroup) | (4-89); 2085F/600M<br>(60-89); 117F/85M | Identification<br>Self-report | Food neophobia<br>questionnaire | Older adults with olfactory dysfunction had a higher level of food neophobia; this was not the case for younger participants with and without OD. |
| (Manesse et al., 2017) | individuals with olfactory dysfunction (N=39)<br>CG (N=40) | 55.72 ± 2.08*; 22F/18M<br>56.82 ± 1.84*; 22F/18M | Identification<br>Self-report | Food questionnaire | Individuals with OD were more neophobic than control participants. |
| (Pelchat, 2000) | Community-dwelling adults;<br>Experiment 1 (N=51)<br>Young adults with NOR (N=16)<br>Older adults with NOR (N=15)<br>Older adults with OD (N=20)<br>Experiment 2 (sauce could not be smelled) (N=54):<br>NOR Young (N=15)<br>NOR Older adults NOR (N=15)<br>Older adults with OD (N=15) | (18-25)<br>Older adults (62-85); 31F/20M<br>Young (19-34)<br>Older adults: (64-85); 28F/17M | Threshold | Four food stimuli, rated on familiarity, willingness to taste, and pleasantness of the food's odor | No significant effect of olfactory dysfunction on willingness to try new food in older adults compared to healthy older adults and healthy young people in both experiments; older adults with OD were significantly more willing to try the unpleasant-smelling foods than the young adults. |
| (Stankovic et al., 2021) | Children with ADHD (N=36)<br>Children without ADHD (N=36) | 14 (13-16); 7F/29M<br>15 (13-16); 18F/18M | Identification;<br>discrimination;<br>threshold | Food Neophobia Test<br>Tool | No correlation between olfactory function and FNTT-score in either group. |
| **Anticipatory food liking** |  |  |  |  |  |
| (de Vries et al., 2018) | Women with breast cancer (N=28)<br>CG (N=28) | 51.0 ± 1<br>51.8 ± 7.6 | Identification;<br>discrimination;<br>threshold<br>Self-report | Macronutrient and Taste<br>Preference Ranking Task | A higher rating of subjective smell function was correlated with a higher liking of low-energy and sweet products. |
| (Fjaeldstad & Smith, 2022) | Normosmic controls (N=166)<br>Individuals with OD (N=522)<br>Olfactory loss (N=271)<br>Parosmia (N=251) | 47 (37-58); 111F/55M<br>47 (34-58); 417F/105M | Self-report | Questionnaire on liking<br>of basic tastants and<br>food items | Liking scores of food items were lower in participants with olfactory dysfunction compared with normosmic controls. |
| (Mattes et al., 1990) | Individuals with a distorted sense of smell (N=60)<br>Individuals with just smell loss (N=58)<br>CG (N=40) | 54.3 ± 16.4; 31F/29M<br>50.0 ± 15.2; 28F/30M<br>51.7 ± 17.8; 30F/10M | Identification;<br>threshold;<br>Self-report | Dietary behavior<br>questionnaire | Changes in food liking were more prominent in individuals with a distorted sense of smell than only smell loss; the most avoided foods in individuals with distortion were sweets and meat. |
| (Mattes & Cowart, 1994) | Individuals with OD (N=389):<br>● No diagnosis (N=48); HYP (N=64); ANS (N=106); DYS (N=30); PH (N=31); Multi-OD (N=31)<br>● CG (N=79) | ● 50.5 ± 15.7 (15-93); 168F/142M<br>● 48.8 ± 18.8 (20-83); 37F/42M | Identification; detection<br>Self-report | Questionnaire on dietary habits | Food dislike was most common in individuals with dysosmia, phantogeusia, and multiple disorders. |
| (Pellegrino et al., 2020) | Anosmic individuals and healthy controls (N=174)<br>USA<br>● ANS (N = 22)<br>● CG (N = 65)<br>Germany<br>● ANS (N = 22)<br>● CG (N = 65) | USA<br>● 55.7 ± 12.5 (36–76); 12F/10M<br>● 34.7 ± 12.2 (21–63); 42F/23M<br>Germany<br>● 55.2 ± 16.4 (18.–82); 19F/12M<br>● 36.5 ± 16.9 (20.0–81.0); 39F/17M | Identification; discrimination; threshold | Food preference ratings for a 15 foods; preference for certain components in foods | Individuals with OD from the USA had a lower overall liking for food stimuli compared to healthy controls, while no such difference was found in the German population. |
| (Postma, Kok, et al., 2020) | Colorectal cancer patients;<br>Longitudinal:<br>● Patients undergoing chemotherapy (N=15)<br>● CG (N=20)<br>Cross-sectional:<br>● T1 (N=20);<br>● T2 (N=20);<br>● T3 (N=20) | Longitudinal:<br>● 66 ± 7.7; 2F/13M<br>● 67 ± 8.8; 6F/14M<br>Cross-sectional:<br>● T1:63 ± 9.1; 10F/10M<br>● T2:65 ± 8.9; 7F/13M<br>● T3:66 ± 4.7; 7F/13M | Identification; discrimination; threshold<br>Self-report | Macronutrient and Taste Preference Ranking Task | No correlation between olfactory function and liking of macronutrients or taste qualities in all groups. |
| (Seo & Hummel, 2009) | ● Young adults with normal olfactory function (N=30)<br>● Older adults with normal olfactory (N=30)<br>● Older adults with OD (N=30) | ● 22.6 ± 2.9 (18-30), 30F<br>● 66.4 ± 3.7 (60-75), 30F<br>● 67.8 ± 4.4 (62-79), 30F | Identification; discrimination; threshold | Rating of the pleasantness of green tea and brewed coffee (both in 3 concentrations) | Variations in the concentration of tea and coffee did not affect the pleasantness of these drinks in older adults with OD. |
| **Food preferences** |  |  |  |  |  |
| (Aschenbrenner et al., 2008) | ● Individuals with OD (N=176)<br>● NOR (N=12);<br>● HYP (N=75);<br>● ANS (N=89) | ● 57.4 ±14.1; 114F/86M | Identification; discrimination; threshold | Questionnaire providing a dietary alterations score (DAS). | After the onset of smell loss, there is a change in taste preferences towards more salty and spicy food. There was no significant difference between the three diagnostic groups. |
| (de Vries et al., 2019) | Advanced oesophagogastric cancer patients (N=15) | ● 61 ± 9.3; 14F/1M | Identification; discrimination; threshold | Macronutrient and Taste Preference Ranking Task | No correlation between olfactory function and food preferences. |
| (Duffy et al., 1995) | Elderly women with high personal functioning (N=80) | ● 76±6 (65-93) | Identification; detection | Survey on food behavior; food preference for 87 foods on a five-point hedonic scale, | The lower olfactory perception was associated with lower preference for foods with a sour/bitter taste or pungency. |
| (Guyot et al., 2021) | Patients undergoing bariatric surgery (N=220) | ● (19 – 68); 197F/23M | Self-report | Self-designed questionnaire on food preferences | Lower preference for cheese among patients with olfactory dysfunction. |
| (Kim et al., 2003) | ● Korean elderly females (N=41)<br>● Young female adults (N=41) | ● $73.0 \pm 1.1$<br>● $24.4 \pm 0.2$ | Detection;<br>threshold | Self-reported changes in dietary habits | Older adults with higher smell thresholds showed changes in sweet and salt preferences with age. |
| (Manesse et al., 2017) | ● Individuals with olfactory dysfunction (N=39)<br>● CG (N=40) | ● $55.72 \pm 2.08^*$; 22F/18M<br>● $56.82 \pm 1.84^*$; 22F/18M | Identification<br>Self-report | Food questionnaire | No significant consequences of dysosmia were found for most aspects of food preferences. |
| (Postma, De Graaf, et al., 2020) | ● Individuals with olfactory dysfunction (N=71)<br>● CG (N=738) | ● $58 \pm 12$ (22–82); 52F/19M<br>● $55 \pm 15$ (19–84); 456F/282M | Self-report | Macronutrient and Taste Preference Ranking Task | Patients with congenital smell loss are more taste (sweet)- or nutrient (fat) oriented. |
| (Postma, Kok, et al., 2020) | Colorectal cancer patients;<br>Longitudinal:<br>● Patients undergoing chemotherapy (N=15)<br>● CG (N=20)<br>Cross-sectional:<br>● T1 (N=20);<br>● T2 (N=20);<br>● T3 (N=20) | ● $66 \pm 7.7$; 2F/13M<br>● $67 \pm 8.8$; 6F/14M<br>● $1:63 \pm 9.1$; 10F/10M<br>● $2:65 \pm 8.9$; 7F/13M<br>● $3:66 \pm 4.7$; 7F/13M | Identification;<br>discrimination;<br>threshold<br>Self-report | Macronutrient and Taste Preference Ranking Task | Food preferences were not correlated to olfactory function. |
| **Appetite and craving** |  |  |  |  |  |
| (Bossola et al., 2011) | Chronic hemodialysis patients (N=110) | ● $64.6 \pm 14.8$; 40F/70M | Self-report | Hemodialysis Study Appetite questionnaire | Changes in smell were significantly higher in patients with a poor/very poor appetite compared to patients with a good/very good appetite. |
| (Chalouhi et al., 2005) | ● Children with CHARGE syndrome (N=14)<br>● CG (N=25) | ● (6-18 years); 8F/6M<br>● (6-13 years); 14 F/11M | Identification;<br>threshold | Questionnaire on child eating difficulty | No relation between olfactory deficits and the severity of feeding disorders (including poor appetite). |
| (Coelho et al., 2021) | Patients with a COVID-19 infection reported change in sense of smell (N=332) | ● $41.57 \pm 13.72$; 258 F/ 63M | Self-report | Questions regarding quality of life | 55% of the patients reported a reduced appetite. |
| (Drareni et al., 2021) | Cancer patients receiving chemotherapy (N=89) | ● $66 \pm 11.7$; 52F/37M | Self-report | Questionnaire on food behavior | No significant effect of chemosensory profile on self-reported appetite. |
| (Essed et al., 2009) | Older adults with:<br>● Normal olfactory + normal gustatory function (N=25),<br>● Normal olfactory + low gustatory function (N=23),<br>● Low olfactory + normal gustatory function (N=34),<br>● Low olfactory + low gustatory function (N=38) | ● $73 \pm 6$; 19F/6M<br>● $70 \pm 7$; 19F/4M<br>● $73 \pm 6$; 23F/11M<br>● $72 \pm 5$; 21F/17M | Identification;<br>detection | Rating desire for soup on a 10-point scale | No effect of flavor enhancement on the desire for soup in any of the groups. |
| (Arganini & Sinesio, 2015) | Independently living Italian older adults (N=99) | ● (65-101 years); 66F/33M | Identification;<br>discrimination;<br>threshold<br>Self-report | Questionnaire on diminished eating pleasure and appetite | Chemosensory impairment does not diminish appetite in independently living older adults. |
| (Ferris & Duffy, 1989) | Individuals with olfactory dysfunction (N=227)<br>● Young (N=104)<br>● Older adults (N=123) | ● (25-45); 50F/54M<br>● (>= 60 years); 61F/62M | Identification;<br>detection | Nutritional evaluation based on diet history, food-intake analysis, and weight record | 66% of individuals with OD reported no change in appetite. Change in appetite was not dependent on age or duration of the OD. Those with a duration of OD > 3 years did not perceive a change in appetite. |
| (Fluitman et al., 2019) | Community-dwelling Dutch older adults (N=673) | ● (57.8±63.1); 345F/328 M | Identification | Question from the Centre for Epidemiologic Studies Depression scale: "In the past week, I did not feel like eating, my appetite was poor." | No association between olfactory function and appetite. |
| (Fluitman et al., 2021) | Community-dwelling Dutch older adults (N=359)<br>● Hyp (N = 66)<br>● Nor (N = 292) | ● (69–77 years); 150F/209M<br>● 75(71–81); 22F/40M;<br>● 72 (69–77);123F/169M | Identification;<br>discrimination;<br>threshold<br>Self-report | 8-item Council of Nutrition Appetite Questionnaire | No association between olfactory function and appetite. |
| (Kim et al., 2003) | ● Korean elderly females (N=41)<br>● Young female adults (N=41) | ● 73.0 ± 1.1<br>● 24.4 ± 0.2 | Detection;<br>threshold | Self-reported changes in dietary habits | 51% of the older adults reported a decreased appetite. |
| (Lechien et al., 2020) | Mild-to-moderate Covid-19 patients (N=417) | ● 36.9 ± 11.4 (19–77); 263F/154M | Identification | The short version of the Questionnaire of Olfactory Disorders-Negative Statements | Patients with anosmia had significantly more loss of appetite than patients with hyposmia or patients with no OD. |
| (Mattes & Cowart, 1994) | Individuals with a chemosensory disorder (N=389)<br>● No-Diagnosis (N=48);<br>● HYP (N=64);<br>● ANS (N=106);<br>● DYS (N=30);<br>● PH (N=31);<br>● Multi-OD (N=31);<br>CG (N=79) | ● 50.5±15.7; 168F/142M<br>● 28F/20M<br>● 32F/32M<br>● 55F/51M<br>● 20F/10M<br>● 21F/10M<br>● 12F/19M<br>● 48.8 ± 18.8 (20-83); 37F/42M | Identification;<br>detection<br>Self-report | Questionnaire on dietary habits | 22% up to 48% of the individuals reported a decrease in appetite after the onset of OD. |
| (Mattes et al., 1990) | ● Individuals with a distorted sense of smell (N=60)<br>● Individuals with smell loss (N=58)<br>● CG (N=40) | ● 54.3 ± 16.4; 31F/29M<br>● 50.0 ± 15.2; 28F/30M<br>● 51.7 ± 17.8; 30F/10M | Identification;<br>threshold<br>Self-report | Dietary behavior questionnaire | 37% of the individuals with a distorted sense of smell reported a decrease in appetite, compared to 22% of individuals with just smell loss. |
| (Nordin et al., 2011) | individuals with smell loss and with nasal polyposis and asthma (N=50)<br>● HYP (N=16)<br>● ANS (N= 34) | ● 50.5 ± 13.1; 21F/29M | Threshold | Questions about consequences of smell loss, QoL, psychological well-being and distress, and coping strategies | 27% of patients reported worsened appetite. |
| (Omlin et al., 2013) | ● Advanced cancer patients (N=52)<br>● CG (N=52) | ● 63 (25-86); 21F/31M<br>● 64 (26-81); 21F/31M | Self-report | NIS checklist (nutrition impact symptoms) | 27% of the patients reported taste and smell alterations reducing their appetite. |
| **Cooking habits** |  |  |  |  |  |
| (Fjaeldstad & Smith, 2022) | Normosmic controls (N=166)<br>Patients with OD (N=522)<br>● ANS (N=271)<br>● PAR (N=251) | ● 47 (37–58); 111F/55M<br>● 47 (34–58); 417F/105M | Self-report | Cooking and Food Provisioning Action Scale | Total CAFPAS score was significantly different between normosmic participants and participants with olfactory dysfunction. However, age and etiology of OD had no effect. Significant differences in cooking habits were found between individuals with smell loss and distorted sense of smell. |
| (Manesse et al., 2017) | ● Individuals with OD (N=39)<br>● CG (N=40) | ● 55.72 ± 2.08*; 22F/18M<br>● 56.82 ± 1.84*; 22F/18M | Identification;<br>Self-report | Food questionnaire | No differences in cooking habits between groups. |
| (Nordin et al., 2011) | Individuals with smell loss, nasal polyposis and asthma (N=50)<br>● HYP (N=16)<br>● ANS (N= 34) | ● (50.5 ± 13.1); 21F/29M | Threshold | Questionnaire on consequences of smell loss | Difficulty in cooking was the most common interference. |
| (Rowan et al., 2019) | ● Patients with chronic rhinosinusitis (N=70) | ● (52.2±17.1); 32F/38M | Identification;<br>discrimination;<br>threshold | Factor 2 of Questionnaire of Olfactory Disorders Negative Statements | Patients with a lower threshold score had a higher chance of impaired eating-related QoL. |

F (Female); M (Male); ADHD (Attention Deficit Hyperactivity Disorder), ANS (Anosmia); CG (Control Group); CAFPAS (Cooking and Food Provisioning Action Scale); CHARGE (coloboma, heart defects, atresia choanae);
DYS (Dysosmia); GD (Gustatory dysfunction);HYP (Hyposmia); Multi-OD (Multiple diagnoses for olfactory dysfunction); NOR (Normosmia); OD (Olfactory dysfunction); PAR (Parosmia); PH (Phantosmia); QoL (Quality of Life); SD (Standard Deviation); SEM (Standard Error of Mean),
\* Reported with SEM instead of SD

**Table 5.** Included articles for the consummatory phase of eating behavior per category: nutritional intake, adherence to dietary guidelines, consumption frequency, experienced liking, food enjoyment, and eating habits.

| Consummatory phase of eating behavior |  |  |  |  |  |
| --- | --- | --- | --- | --- | --- |
|  | Participants |  | Measurement methods |  |  |
| Authors | Sample | Age: mean ± SD (age range);<br>Gender: F/M | Olfactory<br>dysfunction | Eating behavior | Result |
| Nutritional intake |  |  |  |  |  |
| (Aschenbrenner et al., 2008) | Individuals with OD (N=176)<br>● NOR (N=12);<br>● HYP (N=75);<br>● ANS (N=89) | ● 57.4 ± 14.1; 114F/86M | Identification;<br>discrimination;<br>threshold | Specifically designed<br>questionnaire providing a<br>dietary alterations score | 29% of all individuals with OD reported that they eat less since the onset of olfactory dysfunction. |
| (Belqaid et al., 2014) | Patients under investigation for<br>suspected LC (N=215)<br>● LC=117<br>● CG=98 | ● 68 ± 9; 63 F /54 M<br>● 66 ± 10; 43F/ 55M | Self-report | Patient-Generated Subjective Global<br>Assessment | Olfactory dysfunction decreases food intake in cancer patients. |
| (Brisbois et al., 2011) | ● Adult advanced cancer<br>patients (N=192) | ● 64.3 ± 12.4; 95F/97M | Self-report | Three-day dietary record | Chemosensory alterations were associated with decreased caloric and protein intakes. |
| (Duffy et al., 1995) | ● Elderly women with high<br>personal functioning (N=80) | ● 76±6 (65-93 years); | Identification; detection | Food frequency<br>questionnaire, 24-h food<br>records (N=5) | The lower olfactory perception was associated with a higher intake of sweets and a lower intake of low-fat milk products. |
| (Essed et al., 2009) | Older adults with:<br>● Normal olfactory + normal<br>gustatory function (N=25),<br>● Normal olfactory + low<br>gustatory function (N=23),<br>● Low olfactory + normal<br>gustatory function (N=34)<br>● Low olfactory + low<br>gustatory function (N=38) | ● 73 ± 6; 19F/6M<br>● 70 ± 7; 19F/4M<br>● 73 ± 6; 23F/11M<br>● 72 ± 5; 21F/17M | Identification;<br>detection | Soup intake | No significant difference in intake between the plain soup and the flavor-enhanced soup in any of the groups. |
| (Ferris et al., 1985) | ● Lifelong ANS (N=9)<br>● Mid-ANS (N=22)<br>● Onset ANS (N=22)<br>● CG= (N=33) | ● 41.7 ±17.2;3F/6M<br>● 51.6 ± 13.41;3F/9M<br>● 50.2 ±17.5;11F/11M<br>● 49.9 ±16.2;17F/16M | Identification;<br>threshold | Nutrition interview; 24-hour<br>recall, the two-day record of<br>food intake | No differences were found in the nutrient intake between people with OD and the control group. |
| (Ferris & Duffy, 1989) | Individuals with olfactory<br>dysfunction (N=227)<br>● Young (N=104)<br>● Older adults (N=123) | ● (25-45); 50F/54M<br>● (>= 60) 61F/62M | Identification;<br>detection | Nutritional evaluation based<br>on diet history, food-intake<br>analysis, and weight record | 62% reported no change in food intake. |
| (Fluitman et al., 2021) | Community-dwelling Dutch older adults (N=359)<br>● HYP (N = 66);<br>● NOR (N = 292) | ● (69–77);150F/209M<br>● 75(71–81); 22F/40M;<br>● 72(69–7);123F/169M | Identification;<br>discrimination;<br>threshold<br>Self-report | Food frequency<br>questionnaire | People with hyposmia and those with a normal sense of smell had similar total energy intake. Individuals with hyposmia consumed significantly less protein and alcohol and more carbohydrates |
| (Henkin, 2014) | ● Individuals with HYP (N=56)<br>● CG (N=27) | ● (46 ± 2); 33F/23M<br>● 28 ± 2 (19-56); 22F/5M | threshold;<br>rating | Questions on salt usage | In individuals with hyposmia, most of them estimated their salt usage increased on average 2.8 times compared to before their hyposmia onset. |
| (Hutton et al., 2007) | ● Advanced cancer patients (N=66) | ● 65.4±12.4; 36F/30M | Self-report | Three-day dietary records; | Smell complaint scores were inversely related to energy intake and affected the macronutrient composition of the diet (lesser proportion of fat). |
| (Kim et al., 2003) | ● Korean elderly females (N=41)<br>● Young female adults (N=41) | ● 73.0 ± 1.1<br>● 24.4±0.2 | detection;<br>threshold | Food interview; 24 hr-recall | Higher smell threshold results in lowering consumption of meats, eggs, and cereals and lowering calories, protein, fat, carbohydrate, and minerals. |
| (Kong et al., 2016) | South Korean adults<br>● OD (N=1,332)<br>● CG (N=23,158) | ● 14,663F/9,827M<br>● mean age=54.7<br>● mean age=45.9 | Self-report | 24-hour recall | Olfactory dysfunction was associated with reduced fat intake for the whole group and showed differential effects depending on age and gender. |
| (Manesse et al., 2017) | ● Individuals with Dysosmic (N=39)<br>● CG (N=40) | ● 55.72 ± 2.08*; 22F/18M<br>● 56.82 ± 1.84*; 22F/18M | Identification;<br>Self-report | Food questionnaire | Individual with OD Increased use of condiments (sugar, mayonnaise, sour cream) for flavor enhancement |
| (Manesse et al., 2021) | ● French individuals (N=3685)<br>● Older adults (N=202) (subgroup) | ● (4 -89); 2085F/600M<br>● (60-89); 117F/85M | Identification;<br>Self-report | Food questionnaire | Olfactory dysfunction led to greater consumption of dairy products and meat and lower consumption of vegetables in older adults but did not affect food intake in younger participants. |
| (Mattes et al., 1990) | ● Individuals with a distorted sense of smell (N=60)<br>● Individuals with smell loss (N=58)<br>● CG (N=40) | ● 54.3 ± 16.4;31F/29M<br>● 50.0 ± 15.2; 28F/30M<br>● 51.7 ± 17.8; 30F/10M | Identification;<br>detection;<br>Self-report | Food frequency<br>questionnaire; 24-h recall; 2-d food record | Both individuals with a distorted sense of smell and those with just smell loss reported changes in intake (i.e., decreased or increased); Those who reported changes indeed consumed less nutrients. |
| (Mattes & Cowart, 1994) | Patients with a chemosensory disorder (N=389)<br>● No-Diagnosis (N=48);<br>● HYP (N=64);<br>● ANS (N=106);<br>● DYS (N=30);<br>● PH (N=31);<br>● Multi-OD (N=31);<br>● CG (N=79) | ● 50.5±15.7; 168F/142M<br>● 28F/20M<br>● 32F/32M<br>● 55F/51M<br>● 20F/10M<br>● 21F/10M<br>● 12F/19M<br>● 48.8 ± 18.8 (20-83);<br>37F/42M | Identification;<br>detection;<br>Self-report | 3-day food record; | There were significant differences in intake of carbohydrates, dietary fiber, and total sugars between patient groups and healthy controls. Increased use of sugar and salt was reported by 39-57% of the patients. |
| (Merkonidis et al., 2015) | Individuals with chemosensory disorders (N=269) | ● M (52.3 ± 12.3)<br>● F (50.6 ±12.3),<br>● 149 F/116M<br>● Gender N/A (N=4) | Self-report | The questionnaire comprises six sections: eating with a smell and taste disorder | More patients reported eating less (18.6%) than eating more (7.3%). |
| (Omlin et al., 2013) | Advanced cancer patients (N=52)<br>CG (N=52) | 63 (25-86); 21F/31M<br>64 (26-81); 21F/31M | Self-report | NIS checklist (nutrition impact symptoms) | 27% of the patients reported taste and smell alterations reducing their oral intake. |
| (Rasmussen et al., 2018) | Patients with diabetes (N=428)<br>CG (N=2776) | 62.2 (11.2); 193F/235M<br>58.1 (12.2); 231F/197M | Identification | Dietary Interview | Patients with OD had a lower daily caloric intake and a lower intake of carbohydrates and sodium. Controls with OD also had a lower daily caloric and fat intake compared to controls without OD. |
| (Rowan et al., 2019) | Patients with chronic rhinosinusitis (N=70) | 52.2 ± 17.1; 32F/38M | Identification; discrimination; threshold | Questionnaire of Olfactory Disorders Negative Statements | Patients with a lower threshold score had a higher chance of impaired eating-related QoL. |
| (Schubert et al., 2012) | Participants in the Beaver Dam Offspring Study (N=2838) | 49(21–84); 1545F/1293 M | Identification | Dietary choices were assessed by 4 questions. | Overall, olfactory impairment was not linked to the quantity of fruit or vegetable servings or how often salt or sugar was added to food. |
| (Stevenson et al., 2020) | Individuals with olfactory dysfunction (N=222)<br>Congenital (N=10)<br>Idiopathic (N=66)<br>Post-infection (N=63)<br>Sino-nasal disease (N=34)<br>Trauma (N=33)<br>Miscellaneous (N= 17) | 55.6 ± 16.5; 127F/95M | Identification; discrimination; threshold | The 26-item German version of the DFS (dietary fat and sugar scale) to assess intake of discretionary processed foods—a Western-style diet (rich in sugar, salt, and saturated fat) | The etiology-based approach revealed both positive and negative associations between olfactory performance and consumption of a WSD. |
| **Adherence to dietary guidelines** |  |  |  |  |  |
| (Ferris & Duffy, 1989) | Individuals with olfactory dysfunction (N=227)<br>Young (N=104);<br>Elderly (N=123) | (25-45) ;50F/54M<br>(>=60); 61F/62M | Detection; Identification | Nutritional evaluation based on diet history, food-intake analysis, and weight record | Individuals with OD demonstrated a low intake of vitamin B6 and zinc intake was affected by age, gender, severity, and duration of the dysfunction. |
| (Fluitman et al., 2021) | Community-dwelling Dutch older adults (N=359)<br>HYP (N = 66);<br>NOR (N = 292) | 69–77 years; 150F/209M<br>75(71–81); 22F/40M<br>72 (69–77); 123F/169M | Identification; discrimination; threshold<br>Self-report | Food frequency questionnaire | Hyposmics had worse scores for the AHEI and tended to have worse adherence to the MDS. |
| (Gopinath et al., 2016) | Blue Mountains Eye Study participants<br>Older adults with OD (N=89)<br>Older adults without OD (CG) (N = 468) | 70.7 ± 6.5; 305F/252M<br>74.0 (6.7); 34F/55M<br>70.0 (6.3); 371/197M | Identification | Validated semiquantitative, food frequency questionnaire used to calculate adherence to dietary guidelines and total dietary score | Older adults showed a poorer adherence to dietary guidelines when they experienced olfactory dysfunction. |
| (Postma, De Graaf, et al., 2020) | Individuals with olfactory loss (N=105)<br>CG (N=738) | 58 ± 12 (22-82); 77F/28M<br>55 ± 15 (19-84); 457F/281M | Self-report | Adherence to Dutch Dietary Guidelines | Individuals with OD had lower adherence to the dietary guidelines for fiber, trans fatty acids, and alcohol. |
| (Rawal et al., 2021) | Older adults from the NHANES survey<br>OD (N=1399)<br>CG (N=4957) | 58.6 ± 0.4 (SEM)<br>57.7 ± 0.2 (SEM)* | Self-report | 24h dietary recall, used to calculate the Healthy Eating Index score | Individuals with OD have a lower diet quality and consume more foods with higher energy density, and have lower consumption of vegetables. |

| Consumption frequency |  |  |  |  |  |
| --- | --- | --- | --- | --- | --- |
| (Fjaeldstad & Smith, 2022) | individuals with OD (N=522)<br>• Olfactory loss (N=271)<br>• Parosmia (N=251)<br>• Normosmic controls (N=166) | • 47 (34–58); 417F/105M<br>• 47 (37–58); 111F/55M | Self-report | Questionnaire on frequency of intake of basic tastants and food items | The difference in frequency of intake was not significant between individuals with OD and the normosmic controls. |
| (Han et al., 2018) | • Individuals with OD (N=60)<br>• CG (N=60) | • 59.8 ± 12.4; 43F/17M<br>• 58.4 ± 14.6; 38F/22M | Identification; discrimination; threshold | Self-designed questionnaire for Dietary evaluation | Olfactory dysfunction was not associated with changes in the number of meals per day compared to healthy controls. Individuals with OD consumed alcohol less frequently than healthy controls. |
| (Kremer et al., 2014) | Dutch older adults with and without olfactory impairment (N=321)<br>• NOR (N=213)<br>• HYP (N=108) | • 66.2(5.5);124F/89M<br>• 68.3(6.7);47F/61M | Identification; discrimination; threshold | An extensive questionnaire containing various questions on eating behavior | No difference in eating frequency was observed between olfactory-impaired and unimpaired people. |
| (Mattes et al., 1990) | • Individuals with a distorted sense of smell (N=60)<br>• Individuals with smell loss (N=58)<br>• CG (N=40) | • 54.3 ± 16.4; 31F/29M<br>• 50.0 ± 15.2; 28F/30M<br>• 51.7 ± 17.8; 30F/10M | Identification; threshold; Self-report | Food frequency questionnaire, 24-h recall, and 2-d food record | Intake frequency was lower in individuals with a distorted sense of smell compared to those with just smell loss or healthy controls. |
| (Mattes & Cowart, 1994) | Individuals with OD (N=389)<br>• No-Diagnosis (N=48);<br>• HYP (N=64);<br>• ANS (N=106);<br>• DYS (N=30);<br>• PH (N=31);<br>• Multi-OD (N=31);<br>• CG (N=79) | 50.5±15.7; 168F/142M<br>• 28F/20M<br>• 32F/32M<br>• 55F/51M<br>• 20F/10M<br>• 21F/10M<br>• 12F/19M<br>• 48.8 ± 18.8; 37F/42M | Identification<br>Self-report | Questionnaire on dietary habits | 46% up to 77% of the Individuals with OD reported alterations in their eating patterns, mostly changes in eating frequency. |
| Experienced Liking |  |  |  |  |  |
| (Essed et al., 2009) | Older adults with:<br>• Normal olfactory + normal gustatory function (N=25)<br>• Normal olfactory + low gustatory function (N=23)<br>• Low olfactory + normal gustatory function (N=34)<br>• Low olfactory + low gustatory function (N=38) | • 73 ± 6; 19F/6M<br>• 70 ± 7; 19F/4M<br>• 73 ± 6; 23F/11M<br>• 72 ± 5; 21F/17M | Identification; detection | Rating liking of soup on a 10-point scale | No effect of flavor enhancement on liking of soup in any of the groups. |
| (Kremer et al., 2007) | Older adults (N=52)<br>Young CG (N=55) | 71.3 (61–86); 36F/16M<br>22.7 (18–35); 31F/24M | Identification; detection | Liking of custard desserts and tomato drinks after applying flavor enhancement, textural change, and/or irritant addition | Multi-sensory enhancement did not affect increasing food liking in older adults with OD. |
| (Kremer et al., 2014) | Dutch young and older people with and without olfactory impairment (N=122)<br>Young consumers (N=38)<br>HYP old (N=43)<br>NOR old (N=41) | (32.3 ± 8.9); 22F/16M<br>(68.5 ± 5.9); 18F/25M<br>(65.1 ± 5.2); 28F/13M | Identification; discrimination; threshold | Food liking was assessed on a 100 mm horizontal visual analog scale | Older adults with and without OD increased food liking in multi-sensory enrichment in warm meal components. |
| (Novakova et al., 2012) | Individuals with congenital anosmia (N=15)<br>CG (N=15) | 31.0 ± 9.9 (20-42); 3F/2M<br>27.8 ± 5.2 (21-39); 2F/3M | Identification; discrimination; threshold | Rating the pleasantness of a stimulus (banana) during consumption on a 21-point scale | In individuals with congenital anosmia, the decline in pleasantness during consumption was lower than in healthy controls. |
| (Seo & Hummel, 2009) | Young adults with normal olfactory function (N=30)<br>Older adults with normal olfactory (N=30)<br>Older adults with OD (N=30) | 22.6 ± 2.9 (18-30), 30F<br>66.4 ± 3.7 (60-75), 30F<br>67.8 ± 4.4 (62-79), 30F | Identification; discrimination; threshold | Rating of the pleasantness of green tea and brewed coffee (both in 3 concentrations) | Variations in the concentration of tea and coffee did not affect the pleasantness of these drinks in older adults with OD. |
| **Food enjoyment** |  |  |  |  |  |
| (Arganini & Sinesio, 2015) | Independently living Italian older adults (N=99) | (65 -101); 66F/33M | Identification; discrimination; threshold<br>Self-report | Questionnaire on diminished eating pleasure and appetite | Chemosensory impairment does not diminish eating pleasure in independently living older adults. |
| (Coelho et al., 2021) | Patients with a COVID-19 infection reported change in sense of smell (N=332) | 41.57 ±13.72; 258F/63 M | Self-report | Questions regarding the quality of life (QOL) and safety concerns | Reduced enjoyment of food was the most common complaint (87%). |
| (Ferris et al., 1985) | Lifelong ANS (N=9);<br>Midterm-ANS (N=22);<br>Onset ANS (N=22);<br>CG (N=33) | 41.7 ±17.2;3F/6M<br>51.6± 13.41;3/9<br>50.2 ±17.5;11/11<br>49.9 ±16.2;17/16 | Identification; threshold | Score enjoyment between 1-5 | The recent-onset and mid-term anosmics had experienced significantly lower food enjoyment compared to healthy controls, whereas individuals with life-long olfactory dysfunction did not. |
| (Ferris & Duffy, 1989) | Individuals with olfactory dysfunction (N=227)<br>Young (N=104);<br>Older adults (N=123) | (25-45);50F/54M<br>(>=60);61F/62M | Identification; detection | Nutritional evaluation based on diet history, food-intake analysis | In the total sample, food enjoyment was very great (27%) or somewhat (42%) decreased regardless of gender or severity of smell disorder. Among older adults, food enjoyment was only diminished if the duration of OD was < 3 years. |
| (Hutton et al., 2007) | Advanced cancer patients (N=66) | ● 65.4±12.4; 36F/30M | Self-report | Three-day dietary records | Food enjoyment was lower among individuals with severe chemosensory complaints vs. patients with mild or moderate OD. |
| (Liu et al., 2021) | Individuals with smell loss (N=133)<br>● HYP (N=61)<br>● ANS (N=72) | ● 55.9 +- 16.3; 81F/52M | Identification; discrimination; threshold | 19-item Questionnaire of Olfactory Disorders | Self-perceived senses of taste and flavor are more dominantly associated with food enjoyment, in contrast to the patient-perceived sense of smell. |
| (Manesse et al., 2017) | ● Individuals with DYS (N=39)<br>● CG (N=40) | ● 55.72 ± 2.08*; 22F/18M<br>● 56.82 ± 1.84*; 22F/18M | Identification;<br>Self-report | Food questionnaire | Food enjoyment was lower in individuals with quantitative smell disorder |
| (Manesse et al., 2021) | French individuals from the general population (N=3685) | (4 -89); 2085 F/600 M | Identification<br>Self-report | Food neophobia questionnaire | Better smell function was associated with greater food enjoyment. |
| (Mattes et al., 1990) | Individuals with a distorted sense of smell (N=60)<br>● Patients with smell loss (N=58)<br>● CG (N=40) | ● 54.3 ± 16.4; 31F/29M<br>● 50.0 ± 15.2; 28F/30M<br>● 51.7 ± 17.8; 30F/10M | Identification; detection<br>Self-report | Dietary behavior questionnaire | Decreased food enjoyment was higher in individuals with distorted smell than in those with smell loss. |
| (Mattes & Cowart, 1994) | Individuals with OD (N=389)<br>● No diagnosis (N=48);<br>● HYP (N=64);<br>● ANS (N=106);<br>● DYS (N=30);<br>● PH (N=31);<br>● Multi-OD (N=31)<br>CG (N=79) | 50.5 ± 15.7 (15-93);168F/142M<br>● 28F/20M<br>● 32F/32M<br>● 55F/51M<br>● 20F/10M<br>● 21F/10M<br>● 12F/19M<br>48.8 ± 18.8 (20-83); 37F/42M | Identification; detection<br>Self-report | Questionnaire on dietary habits | Most individuals reported decreased food enjoyment; this was highest among people with multiple diagnoses and lowest in those with anosmia. Food enjoyment declined with the duration of OD. |
| (Merkonidis et al., 2015) | Individuals with chemosensory disorders (N=269) | ● M (52.3 ± 12.3)<br>● F (50.6 ±12.3),<br>● 149 F/116M<br>● Gender N/A (N=4) | Self-report | The questionnaire comprises six sections: eating with a smell and taste disorder | The decreased pleasure from eating was the most common complaint. Duration of OD does not affect the declined pleasure. |
| (Nordin et al., 2011) | Individuals with smell loss and with nasal polyposis and asthma (N=50)<br>● HYP (N=16)<br>● ANS (N= 34) | ● 50.5 ± 13.1; 21F/29M | Threshold | Questions about consequences of smell loss;<br>Quality of life questionnaire | 68% of participants had a diminished food enjoyment. |
| (Philpott & Boak, 2014) | Members of the patient support organization Fifth Sense (N=496) | ● 55 (8–95); 318F/178M | Self-report | The survey included questions on the quality of life, depression, and anxiety, impact, and allowed free text entries | People with qualitative disorders have less enjoyment of food in terms of flavor perception, eat unhealthily, and eat less. |
| (Postma, De Graaf, et al., 2020) | Individuals with olfactory loss (N=105) | ● 58±13 (14–87); 73F/32M | Self-report | Questions on food enjoyment | People with OD enjoy eating food less than they did before the onset of their smell loss. |
| (Rowan et al., 2019) | Patients with chronic rhinosinusitis (N=70) | ● (52.2 ±17.1); 32F/38M | Identification; discrimination; threshold | Factor 2 of Questionnaire of Olfactory Disorders Negative Statements | Of the potential effects of their decreased olfaction on QoL, the enjoyment of food and eating was missed the most. |
| (Schubert et al., 2012) | Participants in the Beaver Dam Offspring Study (N=2838) | ● (21–84, mean 49); 1545F/1293 M | Identification | Two questions on the impact of olfactory impairment on food enjoyment | Participants with olfactory impairment were less likely to report that food tasted as good as it used to, or that they experienced food flavors the same. |
| **Eating habits** |  |  |  |  |  |
| (Aschenbrenner et al., 2008) | Individuals with OD (N=176)<br>● NOR (N=12);<br>● HYP (N=75);<br>● ANS (N=89) | ● (57.4±14.1); 114F/86M | Identification; discrimination; threshold | Specifically designed questionnaire providing a dietary alterations score. | Olfactory dysfunction negatively affects social food-related activities |
| (Philpott & Boak, 2014) | ● Members of the patient support organization, Fifth Sense (N=496) | ● 55 (8–95); 318F/178M | Self-report | Survey including questions on the quality of life; depression and anxiety; impact; free text entries | Patients reported avoidance of birthdays, family events, dinner parties, and eating out. |
| (Ferris & Duffy, 1989) | Individuals with OD(N=227)<br>● Young (N=104)<br>● Elderly (N=123) | ● (25-45 years); 50F/54M<br>● (>=60 years); 61F/62M | Identification; detection | Nutritional evaluation based on diet history, food-intake analysis, and weight record | 67% of the individuals reported changes in eating habits. The severity of smell loss has no effect. |
| (Han et al., 2018) | ● Individuals with OD (N=60)<br>● CG (N=60) | ● 59.8± 2.4; 43F/17M<br>58.4 ± 14.6; 38F/22M | Identification; discrimination; threshold | Self-designed questionnaire | No effect of OD on eating habits. |
| (Lechien et al., 2020) | ● Mild-to-moderate Covid-19 patients (N=417) | ● (36.9±1.4); 263F/154M | Self-report | The short version of the Questionnaire of Olfactory Disorders-Negative Statements | Patients with anosmia ate out significantly less than patients with hyposmia or patients with no OD. |
| (Rowan et al., 2019) | ● Patients with chronic rhinosinusitis (N=70) | ● (52.2±17.1); 32F/38M | Identification; discrimination; threshold | Factor 2 of Questionnaire of Olfactory Disorders Negative Statements | Patients with a lower threshold score had a higher chance of impaired eating-related QoL. |

F (Female); M (Male); ANS (Anosmia), CG (Control Group); DYS (Dysosmia); GD (Gustatory dysfunction);HYP (Hyposmia);LG (Lung Cancer); MDS (Mediterranean Diet Score); Multi-OD (Multiple diagnoses for olfactory dysfunction); NOR (Normosmia); OD (Olfactory dysfunction); PAR (Parosmia); PH (Phantosmia); QoL (Quality of Life), SD (Standard Deviation); SEM (Standard Error of Mean); WSD (Western Style Diet).
\* Reported with SEM instead of SD

## 3. Results: the impact of olfactory dysfunction on eating behavior

### 3.1 Study characteristics and design

The characteristics of the 49 studies included in the review are summarized in Table 3 to Table 6. The studies were conducted in multiple countries, including South Korea, The Netherlands, Germany, Italy, France, the USA, and Australia. Most studies were cross-sectional (N=44). Only Gopinath et al. (Gopinath et al., 2016), De Vries et al. (de Vries et al., 2019), (de Vries et al., 2018) and Postma et al. (Postma, Kok, et al., 2020) applied a longitudinal study design; Essed et al. (Essed et al., 2009) used a single-blind, within-subjects, cross-over design. Sample sizes varied between 15 cancer patients (de Vries et al., 2019) and 24,490 participants from a population-based study in Korea (Kong et al., 2016). Overall, study participants, including control groups, were divided into five populations: 16 studies included individuals with general olfactory dysfunction (quantitative as well as qualitative; N=3,175); 11 studies included older adults (N=3,071); 10 studies included cancer patients (N=933); and 8 studies included individuals with various underlying disorders causing the olfactory dysfunction (e.g., Covid-19 (Coelho et al., 2021),(Lechien et al., 2020); CHARGE^1^ syndrome (Chalouhi et al., 2005); or diabetes (Rasmussen et al., 2018), N=4,394). Additionally, there were 4 population-based studies (N=37,369).

### 3.2 Anticipatory phase of eating behavior

This category includes food neophobia, food preferences, (anticipatory) food liking, appetite and craving, and cooking habits. To explore these aspects more precisely and consistently, the following terminology will be employed in this context. *Food neophobia* is the reluctance to try new or unfamiliar foods (Pelchat, 2000). The evaluative attitudes that people express toward foods are referred to as *food preferences* (Styles, 2003); these are based on a variety of intrinsic and extrinsic factors, including personal preferences, cultural influences, nutritional concerns, and health status (Féart et al., 2013).

*(Anticipatory) food liking* refers to the anticipated pleasure (hedonic pleasure) derived from the food or its sensory properties (Finlayson et al., 2012); this is one of the key drivers of food consumption (Wanich et al., 2018). *Appetite* is defined as a person’s desire to search for, select, and consume foods in general (De Graaf et al., 2004). On the other hand, a food *craving* is an intense urge to consume a specific food (Weingarten & Elston, 1990), (Hill et al., 1991). Cooking habits refer to the individual’s practices and routines related to food preparation and cooking. In the context of this study, it includes aspects such as cooking for oneself and any difficulties or changes in cooking behavior due to the condition of olfactory dysfunction. Table 4 shows the articles that were included for the anticipatory phase of eating behavior per category.

#### 3.2.1 Food neophobia

The general population (Manesse et al., 2017) as well as older adults (Manesse et al., 2021) with olfactory dysfunction are more neophobic than healthy controls. In contrast, this was not the case for younger participants (Manesse et al., 2021). In addition, older adults with OD were equally willing to try new foods compared to healthy older adults and healthy young people. However, older adults with OD were significantly more willing to try foods with an unpleasant odor than young adults, likely because they were unable to detect unpleasant aspects of food odor (Pelchat, 2000). Lastly, there was no correlation between olfactory function and food neophobia score in children with and without Attention Deficit Hyperactivity Disorder (ADHD) (Stankovic et al., 2021). To summarize, we found contrasting results on the effect of olfactory dysfunction on food neophobia. As food neophobia is a personality trait and depends on factors such as age (Demattè et al., 2013), genetics (Knaapila et al., 2007), and sensory properties of food, such as texture and visual cues, the large variation in food neophobia among the population may account for these contrasting results.

#### 3.2.2 Food preferences

There was no significant difference in food preferences (i.e., preferences for macronutrients (Postma, De Graaf, et al., 2020); vegetables, fruits, meat, fish, starchy foods, and dairy products (Manesse et al., 2017)) between participants who lost their sense of smell during life and healthy controls (Manesse et al., 2017), (Postma, De Graaf, et al., 2020). However, individuals with acquired (i.e., quantitative and qualitative) olfactory dysfunction experienced a shift in preference from sweet, sour, bitter, and fatty tastes to a preference for salty and spicy tastes (Aschenbrenner et al., 2008). Moreover, etiology affected preference for foods high in fat, and preference for sweet foods differed between individuals with OD and controls (Postma, De Graaf, et al., 2020). Individuals with congenital OD were found to be more taste-oriented when eating compared to those with acquired smell loss (Postma, De Graaf, et al., 2020).

In older adults, lower olfactory function was associated with a change in sweet and salt preferences (Kim et al., 2003) and in older females, to lower preference for food with a sour or bitter taste or pungent taste (Duffy et al., 1995). In contrast, neither colorectal cancer patients (Postma, Kok, et al., 2020) nor patients with advanced esophagogastric cancer (de Vries et al., 2019) showed a correlation between olfactory dysfunction and food preferences. On the other hand, bariatric surgery patients who exhibited olfactory dysfunction were found to have a reduced preference for cheese (Guyot et al., 2021).

Overall, olfactory dysfunction leads to changes in food preferences, likely to compensate for the changed flavor perception. This is most prominently seen in individuals with congenital anosmia, who are more taste (sweet)- or nutrient (fat) oriented than individuals with acquired OD or individuals with a normal sense of smell (Postma, De Graaf, et al., 2020). However, congenital anosmia does not produce noticeably abnormal food preferences (DOTY, 1977). The exact mechanism of the effect of OD on flavor perception and subsequent food preferences needs further investigation. There are contrasting results on the effect of OD itself on taste ability (Stinton et al., 2010), (B. N. Landis et al., 2010), but olfactory and taste dysfunction are often seen in combination, for example in Covid-19 patients (Boscolo-Rizzo et al., 2022) or cancer patients (Spotten et al., 2017). Lastly, individuals with OD have been found to have a reduced sensitivity to the spiciness of the food, as those with anosmia showed reduced oral irritation in response to chili powder (Han et al., 2021) and a decreased sensitivity towards trigeminal stimuli (Hummel et al., 1996), (Frasnelli et al., 2010). This reduced sensitivity may also contribute to the changes in flavor perception (during the consumption phase) that drive future food preferences in these individuals.

#### 3.2.3 Food (anticipatory) liking

The effect of olfactory dysfunction on the liking of food can differ across cultures. A study conducted in the United States indicated that individuals with OD had a lower overall liking for food stimuli than healthy controls, whereas no significant difference was found between groups in a German population (Pellegrino et al., 2020). Also, the nature of the OD affects food liking. In individuals with smell loss and distorted smell changes in food liking were particularly pronounced in individuals within the latter group with sweets and meat being the most disliked food categories (Mattes et al., 1990). In addition, it was observed that food aversions were more frequent among individuals experiencing a distorted sense of smell and those with multiple diagnoses (e.g., hyposmia and phantosmia) (Mattes & Cowart, 1994).

Olfactory dysfunction can affect food liking in cancer patients undergoing chemotherapy. Among women with breast cancer, a lower self-reported sense of smell was correlated with a lower liking of low-energy and sweet products (de Vries et al., 2018). However, in colorectal cancer patients, olfactory (dys)function was not correlated with food liking (Postma, Kok, et al., 2020). Moreover, in individuals with smell loss and distorted sense of smell following a Covid-19 infection, food liking was significantly lower compared to controls (Fjaeldstad & Smith, 2022).

Overall, most studies show that olfactory dysfunction affects anticipatory food liking. Its effect depends on the nature of OD. Furthermore, external factors such as culture (Pellegrino et al., 2020), or treatment in cancer patients (Boltong & Keast, 2012), can also impact the liking of food.

#### 3.2.4 Appetite and Craving

In individuals with OD, the prevalence of decreased appetite ranges from 22-48% (Mattes & Cowart, 1994), (Nordin et al., 2011), (Ferris & Duffy, 1989). Changes were not dependent on age or extent of the dysfunction, and individuals with long-lasting OD (more than three years) did not perceive a change in appetite (Ferris & Duffy, 1989). Moreover, changes in appetite were more common in individuals with a distorted sense of smell (37%) than in individuals with smell loss (22%) (Mattes et al., 1990). Similarly, more individuals with smell distortion (25%) than with smell loss (19%) reported the development of food cravings (Mattes et al., 1990).

Studies in older adults show mixed findings regarding the relation between olfactory function and appetite. While some studies found no impact of olfactory function on appetite (Arganini & Sinesio, 2015), (Fluitman et al., 2021), (Fluitman et al., 2019), one study among Korean elderly women showed that between 51% and 62% of the participants reported a decreased appetite (Kim et al., 2003). Additionally, olfactory function did not affect the desire for plain soup versus flavor-enhanced soup in older adults (Essed et al., 2009).

Among advanced cancer patients, 27% reported diminished appetite due to their smell alterations (Omlin et al., 2013), while. However, another study showed no significant effect of smell alterations on appetite in patients undergoing chemotherapy (Drareni et al., n.d.).

Among Covid-19 patients with OD, 55% reported a reduced appetite (Coelho et al., 2021). However, reduction in appetite was lower in those with anosmia than in those with hyposmia and without OD (Lechien et al., 2020). In chronic hemodialysis patients there were more changes in smell in patients with a poor appetite compared to those with a good appetite (Bossola et al., 2011). Among children with CHARGE syndrome, no relation between olfactory deficits and the severity of feeding disorders (including poor appetite as part of abnormal feeding behavior) was found (Chalouhi et al., 2005).

The results of these studies suggest that the nature of olfactory dysfunction (qualitative vs quantitative) and its duration can lead to differences in appetite. For instance, it is more likely to experience a decrease in appetite in individuals with short-term olfactory dysfunction, while those with long-term olfactory dysfunction may develop coping strategies (Croy et al., 2014). This is reflected in a higher quality of life in individuals with long-term dysfunction (Shu et al., 2011). Furthermore, while more research is needed, it can be speculated that qualitative olfactory disorders, which typically have more significant effects on eating behavior, may lead to food craving.

#### 3.2.5 Cooking habits

Comparison of individuals with OD to healthy controls revealed no differences in cooking habits, i.e., cooking for oneself and eating prepared meals (Manesse et al., 2017). However, another study found that among individuals with smell loss, difficulty in cooking was the most common interference (Nordin et al., 2011). Additionally, individuals with OD after a Covid-19 infection had lower scores on skills and self-efficacy, and attitude related to cooking compared to healthy controls (Fjaeldstad & Smith, 2022). However, individuals’ age and etiology of the OD (i.e., Covid-19 vs non-Covid-19 (long duration)) have no effect (Fjaeldstad & Smith, 2022).

Generally, individuals experiencing OD face challenges while cooking, including a lack of comfort and inspiration in the kitchen and difficulty in preparing new dishes. Individuals with distorted sense of smell have more challenges with respect to individuals with smell loss, such as a reduced confidence in the ability to deal with unexpected results, and a wish for more time for planning meals.

### 3.3 Consummatory phase of eating behavior

Within the consummatory phase, we defined the following categories: nutritional intake, adherence to dietary guidelines, consumption frequency, (experienced) food liking, food enjoyment, and finally eating habits. *Nutritional intake* is the sum of foods and beverages consumed by a person, including carbohydrates, proteins, fats, vitamins, and minerals (Khalid et al., 2014). *Adherence to dietary guidelines* is defined as following the recommendations for daily nutrient intake as set by national or international health organizations (Van Lee et al., 2012). *Consumption frequency* refers to how often food is consumed during a specified time (Tarasuk & Brooker, 1997). We defined the *experienced food liking* as individuals’ actual liking and sensory satisfaction with the taste, flavor, and texture of the food during consumption. *Food enjoyment* is the overall satisfaction and pleasure derived from consuming food (Styles, 2003) including feelings of pleasure, satisfaction, and well-being during the meal. And last but not least, *eating habits* refer to the behaviors and routines individuals have when it comes to consuming food, and the social aspects of eating, such as eating alone or with others (Jastran et al., 2009).

Table 5 shows the articles that were included for the anticipatory phase per category.

#### 3.3.1 Nutritional intake

Most studies demonstrated an effect of olfactory dysfunction on food intake among the general population. In particular, 29% of individuals reported having reduced their intake since the onset of their OD (Aschenbrenner et al., 2008). Furthermore, individuals reporting intake changes consumed fewer nutrients (Mattes et al., 1990). Notably, a higher proportion (18.6%) reported decreased consumption compared to those increasing intake (7.3%) (Merkonidis et al., 2015). In South Korean adults, olfactory dysfunction was associated with lower fat intake (Kong et al., 2016). Moreover, age and gender were identified as factors that can influence the effect of OD on food intake, as young males with olfactory dysfunction were found to consume less protein compared to their healthy counterparts (Kong et al., 2016). Also, significantly lower consumption of carbohydrates, dietary fiber, and total sugars was observed in individuals with OD with multiple diagnoses compared to healthy controls. However, no marked deficiencies or clinically relevant excesses were found (Mattes & Cowart, 1994).

In contrast, other studies revealed that 62% of the individuals with OD did not exhibit any changes in food intake (Ferris & Duffy, 1989). In addition, no significant difference in nutritional status was observed between individuals with anosmia and healthy controls (Ferris et al., 1985) nor was an association found between olfactory impairment and the consumption rate of vegetables or fruit (Schubert et al., 2012). Moreover, no association between a Western-style diet (i.e., high in sugar, salt, and fat) and OD was identified among the study’s population. However, for the separate subgroups for etiology, various associations were observed between olfactory function and consumption of a Western-style diet (Stevenson et al., 2020).

Most studies reported increased usage of condiments (Manesse et al., 2017) and additional foods among individuals with OD: 15% up to 57% reported altered use of spices (Aschenbrenner et al., 2008), (Merkonidis et al., 2015). Individuals with hyposmia reported an average increase of 2.8 times in the use of salt when compared with before their smell loss (Henkin, 2014). However, another study found no association between the frequency of adding sugar or salt to food and olfactory function (Schubert et al., 2012). Furthermore, a slightly higher rate of change in the use of spices was observed among individuals with a distorted sense of smell (43%) compared to those with smell loss (40%) (Mattes et al., 1990).

Older adults with olfactory dysfunction have been observed to experience changes in their nutritional intake. Studies among elderly women revealed that lower olfactory perception was associated with a higher intake of sweets and a lower intake of low-fat milk products, leading to a nutrient intake profile pointing towards a higher risk for cardiac disease (Duffy et al., 1995) and negatively correlated with the consumption of meats, eggs, cereals, and caloric intake, as well as intake of protein, fat, carbohydrates, and minerals (Kim et al., 2003). Older adults with OD were found to choose different snacks than their unimpaired peers (Kremer et al., 2014) and to consume significantly less protein and alcohol, and more carbohydrates than their normosmic counterparts, though their total energy intake was similar (Fluitman et al., 2021). Additionally, older adults with OD tended to consume more dairy products and meat, and fewer vegetables than younger participants (Manesse et al., 2021), and used more spices to enhance appetite and food intake (Kim et al., 2003). Notably, olfactory function did not affect the intake of plain soup versus flavor-enhanced soup (Essed et al., 2009).

Among advanced cancer patients, chemosensory alterations were found to have a negative impact on energy and macronutrient intake, including protein (Brisbois et al., 2011) and fat (Hutton et al., 2007), with 27% of these patients reporting reduced intake (Omlin et al., 2013). Furthermore, lung cancer patients with smell and taste alterations reported significantly lower food intake than those without such alterations (Belqaid et al., 2014). Additionally, diabetic patients with OD had a decreased daily caloric, carbohydrate, sodium, and fat intake when compared with those without OD. In the same study, controls with OD also had a lower daily caloric intake and lower fat intake compared to the control group without OD (Rasmussen et al., 2018).

To summarize, the literature shows that, regardless of etiology, olfactory dysfunction can lead to a decrease in food intake, potentially resulting in inadequate nutrient intake and deficiencies. Individuals with distorted olfactory function reported avoiding eating because food had an unpleasant flavor, while those with only smell loss reported reduced pleasure as the reason for avoiding eating (Mattes et al., 1990). The effects of olfactory dysfunction do not seem to lead to marked deficiencies in the general population, probably because individuals compensate for the effect of OD by the increased use of condiments and additional foods. However, OD may put older adults at risk of inadequate food consumption, including a decrease in intake and less healthy dietary patterns. Contrary to the findings of Essed et al. (Essed et al., 2009), the study by Mathey et al. revealed that flavor enhancement can improve dietary intake in older adults (Mathey, Siebelink, et al., 2001). This indicates that other factors may also influence nutritional intake, as Essed et al. administered the products in a laboratory setting, while Mathey et al. provided meals during lunch at nursing homes.

#### 3.3.2 Adherence to dietary guidelines

Individuals with OD had a lower diet quality than healthy individuals, consuming more foods with higher energy density, such as saturated fats and added sugars, as well as having a lower consumption of vegetables (Rawal et al., 2021). In contrast, another study showed that there was no difference in *total* adherence to dietary guidelines between individuals with OD and healthy controls. However, individuals did have significantly lower adherence to the guidelines for dietary fiber, trans fatty acids, and alcohol, and a better adherence to salt (Postma, De Graaf, et al., 2020). Furthermore, adequate adherence to dietary recommendations was reported, except for a low intake of vitamin B6 and zinc in individuals with OD (Ferris & Duffy, 1989).

Older adults with olfactory dysfunction showed a poorer adherence to dietary guidelines compared to those without OD (Gopinath et al., 2016), (Fluitman et al., 2021). The self-reported poor smell was associated with lower scores on dietary quality indexes (Fluitman et al., 2021) and female individuals with moderate or severe OD had significantly lower adherence to dietary guidelines than those without OD five years later (Gopinath et al., 2016).

Overall, these results show that individuals with OD have lower adherence to (components of) dietary guidelines, leading to a less healthy diet. Factors that influence adherence include etiology (Postma, De Graaf, et al., 2020), duration (Gopinath et al., 2016), (Ferris & Duffy, 1989) and severity (Ferris & Duffy, 1989) of OD, and age and gender (Gopinath et al., 2016), (Ferris & Duffy, 1989), (Rawal et al., 2021). Therefore, tailored advice for people with olfactory dysfunction, taking factors like the age of the individuals and duration of the OD into account, is necessary to improve adherence to dietary guidelines. Such advice can be derived from more generic strategies employed by individuals with OD, such as focusing on food texture or the visual aspects of food (Fjaeldstad & Smith, 2022) (Ferris & Duffy, 1989).

#### 3.3.3 Consumption frequency

Olfactory dysfunction does not necessarily lead to changes in eating frequency among individuals with acquired olfactory dysfunction, as it was not associated with the number of meals per day nor with the average time spent per meal (Han et al., 2018). Moreover, there was no difference in eating frequency among older adults with olfactory dysfunction when compared to older adults with a normal sense of smell, while those with OD did consume less varied meals (Kremer et al., 2014). Moreover, alcohol consumption was less frequent in individuals with OD than in healthy controls (Han et al., 2018). Other research showed that up to 77% of individuals with OD reported alterations in their eating patterns, mostly changes in eating frequency, with the highest incidence seen in those with multiple diagnoses (e.g., combining hyposmia and phantosmia) (Mattes & Cowart, 1994). Additionally, intake frequency was lower in individuals with a distorted sense of smell compared to those with just smell loss or healthy controls (Mattes et al., 1990), while among individuals with OD after aCovid-19 infection no changes in frequency of intake were found (Fjaeldstad & Smith, 2022).

Concluding, these findings indicate that changes in eating frequency are related to the nature of olfactory dysfunction (i.e., quantitative or qualitative), with a lower consumption frequency observed in those with a distorted sense of smell. This is further supported by evidence from recent work among individuals with a distorted sense of smell after a Covid-19 infection, who indicated that distorted food odors were also mostly perceived as unpleasant (Parker et al., 2022).

#### 3.3.4 Experienced liking

The hedonic value of food typically decreases during consumption; however, in individuals with congenital olfactory dysfunction, this decline in pleasantness was lower than in healthy controls (Novakova et al., 2012). In older adults with OD, variations in the concentration of tea and coffee did not affect the pleasantness of these drinks (Seo & Hummel, 2009). While older adults with and without OD increased food liking in response to multi-sensory enrichment in warm meals (i.e., visual and flavor enrichment in mashed potato) (Kremer et al., 2014), no effects on food liking were observed for flavor and texture enhancement in older adults with OD (Essed et al., 2009), (Kremer et al., 2007). However, changes in the texture affected food pleasantness more in older adults with low olfactory ability compared to older adults with a medium or high olfactory ability (Kremer et al., 2007).

Overall, most studies show that olfactory dysfunction affects food liking. However, it can be assumed a gradual decline of olfactory function, which is common in older adults, has a minor effect on the liking of food, while a sudden change in olfactory function is likely to have a greater effect. This is supported by the finding that olfactory function is not associated with nutritional status (Toussaint et al., 2015) or total energy intake (Duffy et al., 1995). Thus, further research should consider other factors that may affect the liking of food in individuals with olfactory dysfunction, such as the heterogeneity of olfactory dysfunction among older adults (Essed et al., 2009).

#### 3.3.5 Food enjoyment

Food enjoyment, and eating-related quality of life, were found to be positively associated with olfactory function (Manesse et al., 2017), (Manesse et al., 2021), (Postma, De Graaf, et al., 2020), (Nordin et al., 2011), (Ferris et al., 1985), (Rowan et al., 2019), (Schubert et al., 2012). While individuals with acquired olfactory dysfunction had significantly lower food enjoyment compared to healthy controls, those with life-long olfactory dysfunction did not (Ferris et al., 1985). Reduced food enjoyment was also observed in 87% of the individuals in a cohort of Covid-19 patients with olfactory dysfunction (Coelho et al., 2021).

While one study found that decreased food enjoyment declined with the duration of OD (Mattes & Cowart, 1994), another study found no effect of duration (Merkonidis et al., 2015). Individuals with multiple diagnoses related to smell (e.g., hyposmia and phantosmia) (Mattes & Cowart, 1994) and patients with qualitative disorders (Mattes et al., 1990), (Philpott & Boak, 2014) reported decreased enjoyment of food more often compared to those solely suffering from smell loss. The difference between individuals with qualitative and quantitative disorders was most pronounced in those with long-term (> 1 year) problems (Mattes et al., 1990). Additionally, olfactory dysfunction was identified to reduce the enjoyment of food in younger cohorts (Ferris & Duffy, 1989), (Merkonidis et al., 2015), regardless of gender or severity of the disorder (Ferris & Duffy, 1989).

The results obtained for older adults were inconsistent. In one study, over 70% of the participants responded that food enjoyment had diminished to some extent or significantly since the emergence of their OD (Ferris & Duffy, 1989), while another study found that only 18% of the individuals reported a decrease in eating pleasure (Arganini & Sinesio, 2015). The decrease in food enjoyment did, however, diminish as OD duration increased (Ferris & Duffy, 1989). In addition, food enjoyment was lower in patients with advanced cancer who had severe chemosensory complaints compared to those with less severe complaints (Hutton et al., 2007).

Together, these results indicate that OD leads to a decline in food enjoyment, in which young individuals are more affected than older adults. The decline in food enjoyment seems to become less pronounced over time, likely due to the development of coping mechanisms (Ferris & Duffy, 1989). Moreover, the nature of olfactory dysfunction is an important factor in food enjoyment. In individuals with a distorted sense of smell, food enjoyment is more severely impacted, possibly because qualitative olfactory dysfunction likely has a greater impact on flavor perception than quantitative olfactory dysfunction. This was further corroborated by recent studies on parosmia in patients after Covid-19 infections (Parker et al., 2022), (Watson et al., 2021). Moreover, the importance of flavor perception in food enjoyment is supported by the finding of Liu et al. (Liu et al., 2021), who found that food enjoyment was more strongly associated with self-perceived taste and flavor than with smell perception. Also, this is further evidenced by the fact that individuals with lifelong olfactory dysfunction did not show reduced food enjoyment (Ferris et al., 1985). These individuals were never able to perceive the flavor of food fully. This suggests that alternative strategies to enhance the enjoyment of the flavor of food, either through coping strategies (e.g., focusing on other sensory aspects of the food) or by innovative food products (e.g., new food designs), are important to increase overall food enjoyment in patients with olfactory dysfunction.

#### 3.3.6 Eating habits

Comparison of individuals with OD to healthy controls revealed no differences in eating habits, such as emptying a food plate although full (Han et al., 2018). However, another study found that 67% of the individuals reported changes in eating habits (Ferris & Duffy, 1989), mostly by younger females or older adults with shorter OD duration. OD negatively affected socially related eating habits, such as going out for dinner (Aschenbrenner et al., 2008), (Lechien et al., 2020), (Philpott & Boak, 2014), (Rowan et al., 2019). Individuals reported employing coping strategies, including emphasizing the social aspects of the meal (Ferris & Duffy, 1989), to cope with their altered flavor perception.

Concluding, these findings suggest that individuals with OD experience the most difficulty with social activities related to eating, impacting their quality of life (Temmel et al., 2002), (Philpott & Boak, 2014). To cope with the altered perception of flavor, individuals can use a variety of strategies, such as eating with family and friends, altering the quality of spices to stimulate the trigeminal sensation, and focusing on the texture, temperature, and visual presentation of the food. These coping strategies could contribute to a better quality of life, as they might improve social interactions during eating.

## 4. Discussion and conclusion

This systematic scoping review with its primary objective of exploring the range and nature of studies investigating the effect of olfactory dysfunction on distinct aspects of eating behavior reveals a crucial need for standardized assessments in this research area. So far, the exact relationship between olfactory dysfunction and eating behavior has been poorly understood. This may partly be attributed to the complexity of olfactory dysfunction, which can affect eating behavior in multiple ways, as well as a lack of harmonized assessment methods for the distinct aspects of eating behavior. Our analysis of existing literature indicates that this variability can range from self-reported questionnaires to more objective measures like psychophysical tests. Such divergence makes it challenging to draw definitive conclusions or compare results across studies. For example, while some studies used validated tools like food frequency questionnaires, others relied on less standardized methods. This inconsistency highlights the urgent need for standardized, validated tools in this research area.

In discussing the impact of olfactory dysfunction on eating behavior, we note that the heterogeneity in study populations and methodologies leads to diverse findings. However, it can be concluded that there are implications for individuals experiencing olfactory dysfunction, of which decreased food enjoyment is the most outstanding. Moreover, our results show that in the anticipatory phase of eating behavior, food preferences and food liking are most affected in people who experienced a sudden change in olfactory function rather than a gradual decline, probably as the latter group adapts to their decreased olfactory function over time. Moreover, changes in odor perception due to olfactory dysfunction alters the perception of food flavors, resulting in a shift of food preferences towards more taste-based preferences, like salty or savory (i.e., umami). Appetite is more likely to be low in individuals with short-term olfactory dysfunction compared to those with long-term changes. This is because individuals with long-term olfactory dysfunction may habituate to their altered sense of smell. Moreover, individuals with olfactory dysfunction associate cooking with a lack of comfort and inspiration and an inability to make new foods successfully.

Subsequently, changes in preferences in the anticipatory phase can affect food intake and adherence to dietary guidelines in the consummatory phase. This is likely only to a limited extent because food intake is not only regulated by sensory perception but also by other factors, including hunger state and eating habits (Bilman et al., 2017). Additionally, eating behavior is more impacted in individuals with a distorted sense of smell than in those with smell loss; this effect is more pronounced over time (Mattes et al., 1990). Moreover, food enjoyment is most affected in people who experienced a sudden change in olfactory function rather than a gradual decline.

The findings of this review reveal that specific characteristics of olfactory dysfunction influence its impact on eating behavior. For instance, the *duration* and *nature* of dysfunction (sudden versus gradual onset, total loss versus distortion of smell) significantly influence eating behavior. However, the lack of standardized measures makes it difficult to systematically compare these impacts or to understand the complex interplay of factors influencing eating behavior in individuals with olfactory dysfunction. The current result shows that in individuals with a distorted sense of smell, pleasantness, food enjoyment, appetite, consumption frequency, and cooking habits (Fjaeldstad & Smith, 2022) are more affected than in individuals with smell loss. This is attributed to the fact that individuals with a distorted sense of smell perceive food odors differently, and in most cases as unpleasant, rather than simply experiencing them as less intense or non-existent (Watson et al., 2021). Moreover, associating an unpleasant smell (e.g., garbage) with a food item (e.g., coffee) creates an unpleasant sensory experience that can disrupt expectations and create a sense of unease or discomfort. This will make it difficult to enjoy the food and will lead to a lack of appetite. Additionally, the presence of a strong, unpleasant smell can be a distraction and make it difficult to focus on the taste of the food itself. A distorted sense of smell has been linked to a lower overall quality of life (Pellegrino et al., 2021), which is also reflected in eating behavior, including but not limited to decreased enjoyment of food and a decreased consumption frequency.

However, the *duration* of olfactory dysfunction emerges as a central determinant. For instance, as time progresses, individuals with a distorted sense of smell experience a more pronounced effect on their eating behavior compared to those with a complete loss of smell (Mattes et al., 1990). Appetite and food enjoyment are low in individuals with short-term olfactory dysfunction, while in individuals who experience long-term OD, the reduction in food enjoyment will diminish over time. Most likely, this is because they develop coping strategies to deal with their olfactory loss or adapt to it. However, it is noteworthy that such coping strategies, such as focusing on the texture, temperature, and visual presentation of the food, are not only adopted by individuals living with long-term olfactory dysfunction (Croy et al., 2014) but also by those with short-term changes, for instance, patients undergoing chemotherapy (Bernhardson et al., 2012).

In most studies with a heterogeneous population, the effect of *etiology* of olfactory dysfunction on eating behavior was not investigated, mostly because subgroups were too small to compare. Further research involving large-scale studies with diverse etiologies might provide a more comprehensive insight into the effect of the etiology of olfactory dysfunction on eating behavior. However, the effect of duration of olfactory dysfunction might overrule the effect of etiology. This is supported by the evidence shown for the differential impact of acquired versus congenital olfactory dysfunction on eating behavior. Individuals with congenital (i.e., lifelong) anosmia tend to focus more on basic tastes during eating (Postma, De Graaf, et al., 2020), do not show lower food enjoyment, and experience less decline in pleasantness during consumption (i.e., experienced liking) compared to those with acquired dysfunction (Ferris et al., 1985) and healthy controls (Novakova et al., 2012). As individuals with congenital OD were never able to fully perceive the flavor of food, they may have developed coping strategies (Bojanowski et al., 2013) and do not know what they are missing; however, they do report difficulties related to their dysfunction, such as an inability to identify spoiled food (Croy et al., 2014). Moreover, it can be debated if it is the etiology of olfactory dysfunction that affects eating behavior. For example, in cancer patients who experience olfactory dysfunction, the disease itself could also influence their eating behavior. However, in most studies, a control group was included, either consisting of patients with no olfactory dysfunction or healthy controls with a normal sense of smell, which increases the likelihood that the effects on eating behavior in these individuals are caused by the olfactory dysfunction itself. However, in future research, this should be a point of attention, and control groups should therefore be carefully chosen.

In the studies included in this review, a wide range of approaches were employed to assess various aspects of eating behavior. Standardized measures, such as food frequency questionnaires and dietary recalls, are routinely utilized and validated methods for assessing dietary intake. However, as these can also be laborious and time-consuming, in some studies, other measures were used as proxies to measure these aspects of eating behavior. Most of the measures employed in studies were based on surveys, including just a few validated instruments, such as the Questionnaire of Olfactory Disorders (Langstaff et al., 2019) and the Macronutrient and Taste Preference Ranking Task (de Bruijn et al., 2017). Yet, most studies relied on self-developed questionnaires, also because of the unavailability of validated instruments suitable to assess the eating behavior in question. For example, Rowan et al (Rowan et al., 2019) utilized quality-of-life eating-related questions due to the absence of a validated instrument to measure individuals’ food enjoyment.

The evaluation of appetite in the context of olfactory dysfunction was addressed in several studies, with some utilizing a single question to assess this aspect of eating behavior. For instance, Fluitman et al. (Fluitman et al., 2019) applied the following question to evaluate appetite: *“In the past week, I did not feel like eating, my appetite was poor”.* It can be debated whether a single question may be sufficiently sensitive to accurately detect changes in appetite, particularly when comparing it to the situation before the onset of olfactory dysfunction. In addition, some studies employed lab-based settings to measure eating behavior, such as having individuals rate their food desire and liking. These results may not be directly applicable to real-world situations. Hence, it is complex to compare results from various studies due to the use of different methods to assess the same eating behavior aspect such as appetite.

Therefore, we suggest developing standardized questionnaires to measure various aspects of eating behavior in individuals with olfactory dysfunction during both anticipatory and consummatory phases of eating behavior. To measure food intake and adherence to the dietary guidelines, already existing methods, such as food frequency questionnaires (Vijay et al., 2020), or digital dietary assessment tools such as Traqq (Lucassen et al., 2023) can be applied. To assess food liking, appetite, and food preferences, questionnaires need to be standardized and validated. These can then serve as the foundation for future research and, if needed, be extended with additional questions to meet the objectives of specific studies, while still allowing for the comparison of results across studies. Moreover, these questionnaires can include measures that are not included in the current research, such as eating rate.

Additionally, articles included in this review demonstrate a wide range of methodologies employed to measure olfactory function. While some studies incorporated psychophysical tests to objectively diagnose olfactory dysfunction, most studies relied on subjective measures, such as self-reported olfactory function through questionnaires (e.g., AHSP: Appetite, Hunger, and Sensory Perception (Mathey, De Jong, et al., 2001)) and self-report ratings, or incorporated one or more questions on olfactory function into an existing questionnaire. Utilizing objective testing when investigating the effect of olfactory dysfunction on eating behavior will improve the accuracy of the prevalence of smell loss, as already has been demonstrated in Covid-19 patients with olfactory dysfunction (Hannum et al., 2020). The emergence of low-cost, at-home tests for olfactory function during the Covid-19 pandemic, such as the SCENTinel (Hunter et al., 2023), further increases the feasibility of objective testing in studies conducted in a home setting. While at-home tests serve as valuable initial screening tools, more comprehensive insights into olfactory function often require a well-established and validated method such as Sniffin Sticks that offers standardized and quantitative assessment across various olfactory dimensions. Researchers seeking an overview of olfactory assessment methods can refer to the comprehensive work of Parma et al., (Parma & Boesveldt, 2022).

Furthermore, the current body of research largely overlooks the long-term adaptations and coping strategies of individuals with olfactory dysfunction. While some studies hint at these aspects, a standardized approach to measure and evaluate these adaptations is missing. Individuals facing olfactory dysfunction often employ creative strategies to address the impact on their eating behavior. For instance, they may enhanced the flavor through the increased use of spices and condiments, while also focusing on other sensory attributes, like temperature and texture (Aschenbrenner et al., 2008), (Ferris & Duffy, 1989), which can be applied in both the anticipatory and consummatory phase. These coping mechanisms play a pivotal role in the eating behavior of people with olfactory dysfunction since eating pleasure is associated with positive health outcomes (Bédard et al., 2021). Therefore, future research is needed to investigate how people with olfactory dysfunction can modify their diets to make eating enjoyable and palatable, despite their olfactory dysfunction. Most changes in the eating behavior of these individuals are related to changes in flavor perception, as smell is a major contributor to flavor. However, taste and trigeminal senses can also be affected in individuals with olfactory dysfunction. Here lay opportunities for multidisciplinary research, including chefs and food designers, to explore the potential of alternative strategies (e.g., through texture and taste) that people with olfactory dysfunction can apply to have a joyful food experience. Furthermore, research could explore how those with olfactory dysfunction can adjust the way they prepare, cook, and serve meals to enhance other senses, such as sound and touch. This can provide valuable input for promoting healthy eating practices, addressing dietary challenges, and improving overall well-being. Finally, research could study how people with olfactory dysfunction can use technology, such as virtual reality (Li & Bailenson, 2017), to enhance the overall experience of eating.

The systematic scoping review underscores the intricate relationship between olfactory dysfunction and its profound influence on eating behavior, especially in the domains of food liking, preferences, and enjoyment. Qualitative smell loss emerges as a significant factor, with the duration and nature of the dysfunction further shaping its impact. There is a pressing need for further investigations using standardized and validated assessment tools to delve deeper into these findings. Significantly, this research also emphasizes the importance of crafting effective interventions to enrich the eating experience for those affected. By addressing and understanding these connections, we pave the way for enhanced interventions and the promotion of healthier eating habits in individuals with olfactory dysfunction.

## Data Availability

All data produced in the present work are contained in the manuscript

## Declaration of interests

This research was funded by an Aspasia grant of the Netherlands Organization for Scientific Research, awarded to SB. If there are other authors, they declare that they have no known competing financial interests or personal relationships that could have appeared to influence the work reported in this paper.

## Appendix 1: Search strategies for the databases

### Ovid MEDLINE

**(Query for initial search)**

((olfact* OR smell OR chemosensory OR odour ) adj5 (disorder OR loss OR deficit OR dysfunction OR changes OR impairment OR decrease OR alter*).ti,ab,au) OR (anosmia OR hyposmia OR parosmia) AND (appetite OR diet* OR food OR eating OR feed* ) adj5 ( habit OR intake OR pattern OR preference OR choice OR enjoyment OR perception OR behavi* OR pleasure OR quality OR consumption).ti,ab,au

**(Query for additional search)**

((olfact* OR smell OR chemosensory OR odour ) adj5 (disorder OR loss OR deficit OR dysfunction OR changes OR impairment OR decrease OR alter*).ti,ab,au) OR (anosmia OR hyposmia OR parosmia) AND (diet* OR food OR eating OR feed* ) adj5 (pleasantness OR liking OR wanting OR neophobia).ti,ab,au

### PsycInfo

**(Query for initial search)**

AB ( ( appetite OR diet* OR food OR eating OR feed* ) N5 ( habit OR intake OR pattern OR preference OR choice OR enjoyment OR perception OR behavi* OR pleasure OR quality OR consumption) ) AND (AB ( ( olfact* OR smell OR chemosensory OR odour ) N5 ( deficit OR disorder OR loss OR dysfunction OR changes OR impairment OR decrease ) ) OR AB ( anosmia OR hyposmia OR parosmia )) TI ( ( appetite OR diet* OR food OR eating OR feed* ) N5 ( habit OR intake OR pattern OR preference OR choice OR enjoyment OR perception OR behavi* OR pleasure OR quality OR consumption) ) AND (TI ( ( olfact* OR smell OR chemosensory OR odour ) N5 ( deficit OR disorder OR loss OR dysfunction OR changes OR impairment OR decrease ) ) OR TI ( anosmia OR hyposmia OR parosmia ))

**(Query for additional search)**

AB ( ( diet* OR food OR eating OR feed* ) N5 (pleasantness OR liking OR wanting Or neophobia ) ) AND (AB ( ( olfact* OR smell OR chemosensory OR odour ) N5 ( deficit OR disorder OR loss OR dysfunction OR changes OR impairment OR decrease ) ) OR AB ( anosmia OR hyposmia OR parosmia )) TI ( (diet* OR food OR eating OR feed* ) N5 (pleasantness OR liking OR wanting Or neophobia) ) AND (TI ( ( olfact* OR smell OR chemosensory OR odour ) N5 ( deficit OR disorder OR loss OR dysfunction OR changes OR impairment OR decrease ) ) OR TI ( anosmia OR hyposmia OR parosmia ))

### Pubmed

**(Query for initial search)**

(“feeding behaviour“[MeSH Terms] OR eating behav*[Text Word] OR food enjoyment [text word] OR appetite[text word] OR food consumption OR food intake) AND ((“smell“[MeSH] OR olfact*[Text Word] OR chemosensory[Text Word] OR odour[text word]) AND (“physiopathology“[Subheading] OR dysfunction[Text Word] OR deficit[Text Word]) OR (“Anosmia“[Mesh] OR parosmia[Text word]))

**(Query for additional search)**

( “food neophobia” [Text Word] OR “food pleasantness” [Text Word] OR “food wanting” [Text Word] OR “food liking” [Text Word] ) AND ((“smell“[MeSH] OR olfact*[Text Word] OR chemosensory[Text Word] OR odour[text word]) AND (“physiopathology“[Subheading] OR dysfunction[Text Word] OR deficit[Text Word]) OR (“Anosmia“[Mesh] OR parosmia[Text word]))

### Scopus

**(Query for initial search)**

( TITLE-ABS-KEY ( ( olfact* OR smell OR chemosensory OR odour ) W/5 ( disorder OR loss OR dysfunction OR changes OR deficit OR impairment OR decrease OR alter* ) ) OR TITLE-ABS-KEY ( (anosmia OR ypos a OR parosmia ) ) AND T LE-A -KEY ( ( appetite OR diet* OR food OR eating OR feed*) W/5 habit OR intake OR pattern OR preference OR choice OR enjoyment OR perception OR beha* OR pleasure OR quality OR consumption) )

**(Query for additional search)**

(TITLE-ABS-KEY(( olfact* OR smell OR chemosensory OR odour W/5 disorder OR loss OR dysfunction OR changes OR deficit OR impairment OR decrease OR alter* ) OR ( anosmia OR hyposmia OR parosmia )) AND TITLE-ABS-KEY((diet* OR food OR eating OR feed* W/5 lik* OR want* OR pleasantness OR neophobia )))

## Appendix 2: Quality assessment

**Table A2.1.** Quality Assessment Tool for Observational Cohort and Cross-Sectional Studies.

|  |  |
| --- | --- |
| 1 | Was the research question or objective in this paper clearly stated? |
| 2 | Was the study population clearly specified and defined? |
| 3 | Was the participation rate of eligible persons at least 50%? |
| 4 | Were all the subjects selected or recruited from the same or similar populations (including the same time period)? Were inclusion and exclusion criteria for being in the study prespecified and applied uniformly to all participants? |
| 5 | Was a sample size justification, power description, or variance and effect estimates provided? |
| 6 | For the analyses in this paper, were the exposure(s) of interest measured prior to the outcome(s) being measured? |
| 7 | Was the timeframe sufficient so that one could reasonably expect to see an association between exposure and outcome if it existed? |
| 8 | For exposures that can vary in amount or level, did the study examine different levels of the exposure as related to the outcome (e.g., categories of exposure, or exposure measured as continuous variable)? |
| 9 | Were the exposure measures (independent variables) clearly defined, valid, reliable, and implemented consistently across all study participants? |
| 10 | Was the exposure(s) assessed more than once over time? |
| 11 | Were the outcome measures (dependent variables) clearly defined, valid, reliable, and implemented consistently across all study participants? |
| 12 | Were the outcome assessors blinded to the exposure status of participants? |
| 13 | Was loss to follow-up after baseline 20% or less? |
| 14 | Were key potential confounding variables measured and adjusted statistically for their impact on the relationship between exposure(s) and outcome(s)? |

**Table A2.2.** Rating of the manuscripts.

| Group | Good | Fair | Poor |
| --- | --- | --- | --- |
| Older adults | 10 | 1 | 1 |
| Cancer patients | 6 | 4 | 0 |
| Underlying disorders | 4 | 4 | 0 |
| General olfactory dysfunction | 8 | 8 | 0 |
| Population-based studies | 3 | 1 | 0 |
| **Total** | **31** | **18** | **1** |

## Appendix 3: Detailed overview of olfactory function measures

**Table A3.1.** Overview of olfactory function measurements used in the articles included in the review.

| Olfactory outcome measurement | Articles | Category |
| --- | --- | --- |
| **Objective Measures** |  |  |
| Sniffin' Sticks | (Aschenbrenner et al., 2008), (Hannum et al., 2020), (de Vries et al., 2019), (de Vries et al., 2018), (Postma, Kok, et al., 2020), (Vad Andersen & Hyldig, 2015), (Pellegrino et al., 2020), (Demattè et al., 2013), (Knaapila et al., 2007), (DOTY, 1977), (Mattes et al., 1990), (Kremer et al., 2007), (Bossola et al., 2011), (Schubert et al., 2012), (Belqaid et al., 2014) | Identification; discrimination; threshold |
| Odour identification task (OIT) | (Manesse et al., 2021), (Stankovic et al., 2021), (Guyot et al., 2021), (B. N. Landis et al., 2010), (Fjaeldstad & Smith, 2022) | Identification |
| San Diego Odour Identification Test (SDOIT) | (Gopinath et al., 2016), (Schubert et al., 2012) | Identification |
| Smell Identification Test (SIT) | (Manesse et al., 2017) | Identification |
| University of Pennsylvania Smell Identification Test (UPSIT) | (Shu et al., 2011) | Identification |
| Scratch-and-sniff (UPSIT) | (Stinton et al., 2010) | Identification |
| Smell component of the National Health and Nutrition Examination Survey (NHANES) | (Drareni et al., 2021) | Identification |
| 4-Item NHANES Pocket Smell Test (PST) | (Rasmussen et al., 2018) | Identification |
| European Test of Olfactory Capabilities (ETOC) | (Essed et al., 2009), (Postma, De Graaf, et al., 2020) | Detection; identification |
| Odour detection threshold (ODT) | (Manesse et al., 2021), (Stankovic et al., 2021), (Guyot et al., 2021), (Fjaeldstad & Smith, 2022), (Boltong & Keast, 2012), (Brisbois et al., 2011) | Detection |
| French Biolfact olfactory test (olfactory threshold and odor identification) | (Chalouhi et al., 2005) | Identification; threshold |
| Butanol threshold test (BTT) | (Ferris et al., 1985) | Identification; threshold |
| Connecticut Chemosensory Clinical Research Center olfactory test (CCCRC-OT) | (Nordin et al., 2011) | Threshold |
| **Subjective Measures** |  |  |
| Self-Reported Olfactory Loss (self-reported OL) | (Hannum et al., 2020), (Postma, Kok, et al., 2020), [30], (Manesse et al., 2021), (Stankovic et al., 2021), (Stinton et al., 2010), (Spotten et al., 2017), (Han et al., 2021), (Frasnelli et al., 2010), (Novakova et al., 2012), (Pellegrino et al., 2020), (Mattes et al., 1990), (Mattes & Cowart, 1994), , (Bossola et al., 2011), (Ferris et al., 1985), (Hutton et al., 2007), (Van Lee et al., 2012), (Tarasuk & Brooker, 1997), (Henkin, 2014) | Self-report |
| Ratings of odour intensity, pleasantness, familiarity, irritation, and edibility (O_IPFIE) | (Spotten et al., 2017) |  |
| Individual self-reported smell perception scores (VAS) | (de Vries et al., 2018), (Fjaeldstad & Smith, 2022) |  |
| Taste and Smell Survey (TSS) | (Brisbois et al., 2011), (Belqaid et al., 2014) |  |
| CITAS scale (Chemotherapy-induced Taste Alterations) | (Drareni et al., 2021) |  |

## Declaration of interests

☒ The authors declare that they have no known competing financial interests or personal relationships that could have appeared to influence the work reported in this paper.

☐ The author is an Editorial Board Member/Editor-in-Chief/Associate Editor/Guest Editor for *[Journal name]* and was not involved in the editorial review or the decision to publish this article.

☒ The authors declare the following financial interests/personal relationships which may be considered as potential competing interests:

This research was funded by an Aspasia grant of the Netherlands Organization for Scientific Research, awarded to SB. If there are other authors, declare that they have no known competing financial interests or personal relationships that could have appeared to influence the work reported in this paper.

## Credit Author statement

Conceptualization: PP, SB, EP; Data curation: PP, EP; Formal Analysis: PP, EP; Funding acquisition: SB; Investigation: PP, EP; Methodology: PP, SB, EP; Project administration: PP; Resources: PP, EP; Software: PP, EP; Supervision: SB; Validation: PP, SB, EP; Visualization: PP, EP; Writing - original draft: PP, EP; Writing - review & editing: PP, SB, EP

## Footnotes

1 CHARGE is an abbreviation for several of the features common in the disorder: coloboma, heart defects, atresia choanae, growth retardation, genital abnormalities, and ear abnormalities.

## Notes

### Competing Interest Statement

The authors have declared no competing interest.

